# Ability to Detect Changes and Minimal Important Difference of Real-World Digital Mobility Outcomes in Proximal Femoral Fracture Patients

**DOI:** 10.64898/2026.03.06.26347770

**Authors:** C-P Jansen, J Braun, P Alvarez, MA Berge, H Blain, J Buekers, B Caulfield, A Cereatti, S Del Din, J Garcia-Aymerich, JL Helbostad, J Klenk, S Koch, E Murauer, A Polhemus, L Rochester, B Vereijken, MA Puhan, C Becker, A Frei

**Affiliations:** Geriatric Center, Medical Faculty Heidelberg, Heidelberg University, Heidelberg, Germany; Department of Geriatrics, Robert Bosch Hospital, Stuttgart, Germany; Epidemiology, Biostatistics and Prevention Institute (EBPI), University of Zurich (UZH), Zurich, Switzerland; ISGlobal, Barcelona, Spain; Department of Neuromedicine and Movement Science, Norwegian University of Science and Technology, Trondheim, Norway; Department of Geriatric Medicine, St. Olavs hospital, Trondheim University Hospital, Trondheim, Norway; Department of Geriatrics, Montpellier University Hospital and MUSE, Montpellier, France; Insight Centre for Data Analytics, O’Brien Science Centre, University College Dublin, Dublin, Ireland; UCD School of Public Health, Physiotherapy and Sports Science, University College Dublin, Dublin, Ireland; Politecnico di Torino, Department of Electronics and Telecommunications, Torino, Italy; Newcastle NIHR BRC, Translational and Clinical Research Institute, Newcastle University, Newcastle upon Tyne, UK; Universitat Pompeu Fabra (UPF), Barcelona, Spain; CIBER Epidemiología y Salud Pública (CIBERESP); Institute of Epidemiology and Medical Biometry, Ulm University, Ulm, Germany; Study Center Stuttgart, IB University of Health and Social Sciences, Stuttgart, Germany; Uni Basel – University of Basel Department of Sport Exercise and Health; Translational and Clinical Research Institute, Faculty of Medical Sciences, Newcastle University, Newcastle upon Tyne, UK; National Institute for Health and Care Research (NIHR) Newcastle Biomedical Research Centre (BRC), Newcastle University

**Author notes:** Corresponding author Dr. Carl-Philipp Jansen; fax n.a. Equal contributions (C-P Jansen, J Braun).

**Keywords:** Minimal Important Difference, Mobility, Digital Mobility Outcomes, Proximal Femoral Fracture

## Abstract

**Background:** Older adults’ walking has so far been evaluated using standardised assessments of walking capacity within a clinical setting. By taking the evaluation out of the laboratory into the real world, this study provides first evidence of the ability of Digital Mobility Outcomes (DMOs) to detect changes over time and the Minimal Important Difference (MID) in patients after proximal femoral fracture (PFF). This will guide the implementation of DMOs in research and clinical care.

**Methods:** For this multicenter prospective cohort study, 381 community-dwelling older adults were included within one year after sustaining a PFF and assessed at two time points, separated by six months. Walking activity and gait DMOs were measured using a single wearable device worn on the lower back for up to seven days. A global impression of change question and three mobility-related outcome measures (Late-Life Function and Disability Instrument; Short Physical Performance Battery; 4m gait speed) were used as anchor variables. To assess each DMO’s ability to detect changes, we calculated the standardized mean change as effect size. For estimating MIDs, both distribution-based and anchor-based methods were applied, followed by triangulation by experts if at least three anchor-based estimates were available per DMO, resulting in single-point estimates.

**Results:** All three anchor variables demonstrated substantial changes. Overall, 10 out of 24 available DMOs showed large and 7 DMOs moderate positive effects in the expected direction of the respective anchors. Seven DMOs showed no or only small effects. For 12 DMOs, at least three anchor-based estimates were available, enabling MID triangulation. MIDs for walking activity DMOs per day were: a walking duration of 10 minutes, a step count of 1,000 steps, 50 walking bouts (WB), and 15 WBs in WBs over 10 seconds. For gait DMOs, depending on the walking bout length, MIDs for walking speed were between 0.04 m/s and 0.08 m/s, and MIDs for cadence between 4 and 6 steps/minute. Almost all DMOs showed a strong ability to detect improvement in mobility, but rarely in detecting decline.

**Conclusions:** For the first time, MIDs are presented for real-world DMOs in PFF patients. These MIDs inform sample size requirements and interpretation of intervention effects for clinical trials, thereby providing guidance and reassurance for clinicians and regulatory bodies.

## Introduction

### Real-world mobility: the regulatory perspective

The development and evaluation of novel interventions for sarcopenia and frailty require robust measurement of its core domains, among which is walking speed. To be acceptable by regulatory bodies such as the European Medicines Agency (EMA) and the U.S. Food and Drug Administration (FDA), it is imperative that measurements and data are technically valid, supported by experts in the field, have face validity, and are meaningful for patient (1). The SPRINTT and Mobilise-D IMI consortia were set up against this goal. SPRINTT delivered physical capacity data from an exercise intervention (2). The Mobilise-D consortium (3, 4) was inspired by several Phase II trials to test new pharmacological interventions targeting the mTOR pathway and myostatin inhibition to search for anabolic and anti-catabolic approaches to prevent muscle loss in periods of immobilisation and/or to build up muscles to treat sarcopenia. Some of these trials focused on hip fracture patients as a large group faced with both problems. The Mobilise-D consortium set out to address this knowledge gap and to work on a new digital framework to measure real-world mobility over a longer period of time using a single wearable device.

### Real-world mobility: the scientific and clinical perspective

To define mobility, we refer to the outcome of the respective interaction between the Mobilise-D consortium and the European Medicines Agency (EMA), stating that physical mobility is “the ability to move freely and easily without a vehicle” (3). Walking is a key component of mobility that involves musculoskeletal, neurological, cardiorespiratory, sensory, and neuromotor functions (5), and hence is affected by ageing-associated processes, diseases and injuries. Walking has gained a lot of attention in research on healthy aging, and walking speed has been discussed as a “vital sign” – or maybe better a sign of vitality – of health (6). There is clear evidence showing that walking is related to many health outcomes (7, 8). This is especially true for individuals suffering from diseases and injuries that negatively affect mobility such as proximal femoral fracture (PFF). These fractures are a major health issue, and globally the number of hip fractures is expected to rise to 6 million annually by 2050 (9). Most often being caused by a fall (10), PFF are the most detrimental fragility bone fractures that entail severe limitations in mobility and daily functioning (11), from which less than half of the patients can recover (12). Mobility and especially walking ability is a major priority of almost all hip fracture patients (6, 7). It has been shown to predict future incident PFF risk in women (13), but also to be indicative of long-term survivorship overall (14). Consequently, a thorough and in-depth understanding of a patient’s ability to walk is of high importance. This includes the adoption of mobility outcomes as clinical endpoints in intervention studies to mitigate walking disability in patients with pathological gait and loss of function.

Historically, older adults’ walking was evaluated by means of standardised walking capacity assessments in a clinical or laboratory setting. Through recent advancements in movement sensor technology, walking can now be assessed unsupervised and with high ecologically validity in daily life, which has also been shown in PFF patients (15, 16). Another advantage is the possibility to analyse walking beyond snapshot views of single assessments, and instead to deliver continuous data on real-world mobility and walking with higher sensitivity to subtle changes (17). However, inconsistent testing procedures and data evaluation methods have led to scattered approaches and prevented wide-spread adoption of real-world mobility endpoints (18). As part of the Mobilise-D Clinical Validation Study (15), a processing pipeline was validated clinically after having been validated technically (15, 19), leading to a comprehensive list of real-world walking activities and gait parameters, so-called Digital Mobility Outcomes (DMOs), based on data from a single wearable device. In order to evaluate changes in mobility over time in clinical cohorts – including the natural course and in response to a treatment – two central properties are needed: the ability to detect change over time and the Minimal Important Difference (MID). These are key to design studies, to select responsive DMOs, and to calculate sample size requirements. Generally, the MID is used to facilitate the interpretation of patient-reported outcome measures (20). The MID concept states that rehabilitation or treatment results must be larger than the outcome’s MID in order to be of relevance (21).

Against this background, the aim of this study was to provide evidence on the DMOs’ ability to detect changes over time and to estimate the MID of DMOs in patients after PFF.

## Methods

### Study design and participants

In this multicentre, prospective cohort study we used data from the first (study start) and second (6-month follow-up) study visit of the PFF cohort of the Mobilise-D Clinical Validation Study (https://www.mobilise-d.eu/). The full study protocol was previously published (15). All participants gave written informed consent. People with PFF were recruited across sites in Norway (Trondheim and Oslo), Germany (Stuttgart and Heidelberg) and France (Montpellier). Participants were included if they were at least 45 years old, had a surgical treatment for a low-energy fracture of the proximal femur in the past year (that is, patients were included up to 365 days post-surgery), were able to walk 4 meters independently and to consent and comply with study procedures, were willing to wear a single wearable sensor, and able to read and write in the first language of the respective country. Participants were excluded if they were living in a care/nursing home or not able to walk before the hip fracture, and in case of several severe medical circumstances (more information in the study protocol (15)).

During the first visit and 6-month follow-up visit, participants completed interviews and self-administered validated questionnaires and conducted standardised tests. After the visit, they wore a single wearable device on the lower back for seven days: either the McRoberts MoveMonitor+ (McRoberts B.V., Den Haag, the Netherlands; body-worn using a belt) or the Axivity AX6 (Axivity Ltd, Newcastle Upon Tyne, UK; body-fixed using a custom-designed adhesive patch).

The study obtained ethical approval from all relevant Ethical Committees (EC of the Medical Faculty of Eberhard-Karls-University Tübingen [Stuttgart; vote 976/2020BO2], Regional Committee for Medical and Health Professional Research Ethics [Trondheim; vote 216069], Committee of the Protection of Persons, South-Mediterranean II [Montpellier; vote 221 B08]) and was registered at the ISRCTN registry on 12/10/2020 (ISRCTN Number: 12051706).

### Digital Mobility Outcomes

DMOs were calculated on a walking bout (WB) level by running the raw inertial data through the validated Mobilise-D processing pipeline (16, 19, 22). We only considered patients with at least three days of at least 12 hours of measurement during waking time (between 7am and 10pm) (23). Bout level DMOs were aggregated according to the protocol outlined in Koch et al. (24), resulting in the following weekly aggregated DMOs for different walking domains: two DMOs for walking activity amount (daily walking duration and walking bout (WB) step count); seven DMOs for walking activity pattern (e.g., number of WBs/day of different durations); six DMOs for gait pace (e.g., walking speed in WBs longer than 30s); five DMOs for gait rhythm (e.g., cadence in longer WBs); four DMOs for gait bout-to-bout variability (e.g., stride length variability in longer WBs) (see detailed list of all DMOs in Supplementary Material S1).

### Outcomes used as anchors

To assess the DMOs’ ability to detect changes and to estimate the MIDs, we used a global impression of change (GIC) question and three mobility-related outcome measures relevant for older adults after PFF that satisfy the following selection criteria: 1) the outcomes had to have an established MID, and 2) they were accepted as anchors and outcome measures by both the FDA and EMA.

#### Global Impression of change (GIC) question

The GIC question was developed in English following FDA guidance and translated into subsequent destination languages following guidance of the World Health Organization and the ISPOR Task Force for Translation and Cultural Adaptation (25). During the 6-month follow-up visit, the participants were asked “Compared to your first Mobilise-D study visit, how was your walking overall during the last week?”, with answer options “much worse, a little worse, no change, a little better, or much better”.

#### Late-Life Function and Disability Instrument (LLFDI), functional component

The LLFDI was developed as a comprehensive interviewer-administered questionnaire assessing function and disability for use in community-dwelling older adults (26). We used the function component (LLFDI-FC) as an anchor that assesses function in 32 physical activities in three dimensions (upper extremity, basic lower extremity, and advanced lower extremity) with an established MID of 2 (small change) and 5 (substantial change), respectively (27). Acute patients (n=67) were excluded from this analysis since LLFDI questions in this group related to their pre-fracture status, making change estimates invalid.

#### Short Physical Performance Battery (SPPB)

The SPPB assesses lower extremity function and mobility, consisting of three static balance tests, a five-repetition chair-rise test, and a 4-meter walk test. We used the total SPPB score with a MID of 1 point (28) and the 4-meter walk test (supervised gait speed) with a MID of 0.05 m/s (small) and 0.1 m/s (substantial) (29).

### Statistical methods

For all analyses in this paper, release 7.2 (October 2025) of the Mobilise-D data was used. For descriptive statistics, both means with standard deviations and medians with the 25^th^ and 75 ^th^ percentiles (denoted as P25-P75) for continuous data are presented. Categorical data is described using numbers and percentages. All analyses were done separately for each DMO.

In order to investigate the DMOs’ ability to detect changes, we stratified the DMO data according to the participants’ responses to the GIC question and the changes in the three anchor outcomes from the first visit to 6-month follow-up according to the outcomes’ established MIDs (categorized as worse, no change, better). For each of these strata, we calculated mean change, standard deviation, and the standardized mean change as a measure for the effect size. Absolute values between 0.2 and <0.5 were set to be interpreted as small, between 0.5 and <0.8 as medium and ≥0.8 as a large effect (30). For most DMOs, a positive change was interpreted as “better”, except for the two “Stride duration” DMOs (positive change = “worse”). The *a priori* interpretation of change for the five bout-to-bout variability DMOs was less evident, and we did not formulate any well-founded hypotheses for these measures.

For the calculation of suggested MID values, we employed both distribution-based and anchor-based methods (31, 32). For the distribution-based approach, we calculated half of the standard deviation of the DMO at first visit (empirical rule effect size). Anchor-based methods included linear regression and area under the receiver operating curve (ROC). We first calculated Spearman’s correlations for each combination of the changes in a potential anchor and the changes in a specific DMO. To qualify for further investigation and the calculation of a MID value, we predefined that the correlation between these change scores must be ≥0.3. After performing a visual check of linearity between the change scores, we calculated the predicted value of the DMO for the anchor’s MID by using a linear model with the change score of the DMO as the dependent variable and the change in the anchor as the independent variable. We applied the same model for the GIC variable; here we tested the predicted value of the DMO for the stratum “little better”. For the approach based on Receiver Operating Characteristics (ROC), we dichotomized the change of the anchor in two groups (participants whose change exceeded the MID and those not) and we calculated the ROC curves. The MID of the DMO was selected from this curve according to the formula *min*{(*1-sensitivity*)^2^ + (*1-specificity*) ^2^) (33) which defines the point in the top left corner of a ROC curve that differentiates best between the diseased and not-diseased. Individual MID estimates were considered valid if the area under the curve was ≥0.7.

We triangulated MIDs for DMOs if at least three anchor-based estimates were available. The triangulation process included three steps: 1) assessment of the results independently by 11 experts from the fields of geriatrics, orthopedic surgery, movement science, physiotherapy, psychology, biostatistics, and epidemiology in the triangulation process to propose a single MID point-estimate for each DMO (see criteria below); 2) conduct of two independent consensus meetings (University of Zurich team: methodological experts; PFF team: disease-specific researchers and clinical experts, that is, geriatricians, physiotherapists, movement scientists, and orthopedic surgeons) to agree on a single point-estimate; and 3) conduct of a consensus meeting with seven experts of both teams to agree on final point-estimates on July 10 ^th^, 2025. The experts systematically considered the following a priori assessment criteria for their evaluation: Robustness of methods (ROC analyses as most robust, followed by linear regression with outcomes and linear regression with the GIC question [due to potential recall bias]), distribution-based methods deemed to be little informative; relevance of the anchor content (anchors directly related to walking were considered more robust indicators of DMOs’ MIDs than those indirectly related to walking); and number of changers from which the estimates were derived (estimates based on analyses with few persons who changed above/below MID were interpreted with caution).

## Results

### Participant and measurement characteristics

Of the 506 participants PFF patients with valid DMOs at the first visit, 381 had at least one DMO and one anchor measurement at both time points and were therefore included in these analyses (characteristics of entire sample in Table S1). Reasons for exclusion were mainly related to technical and health issues or unavailability for follow-up measurement. Of those, 182 (47.9%) individuals were recruited in Norway, 180 (47.4%) in Germany and 18 (4.7%) in France. Most of the participants were female (n=248, 65.3%), the mean age was 77.3 years, and the median (P25, P75) time between surgery and inclusion in the study was 63 (24, 141) days.

Table 1 shows the characteristics of the participants at first visit (the characteristics of all PFF cohort including also the 125 excluded patients are presented in Table S1).

**Table 1.**
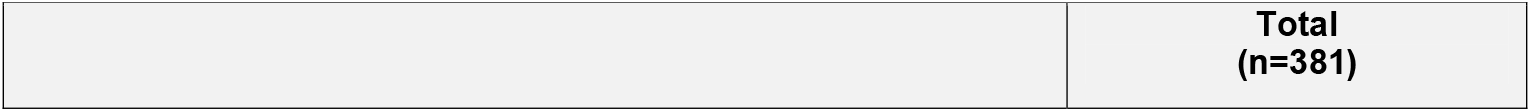
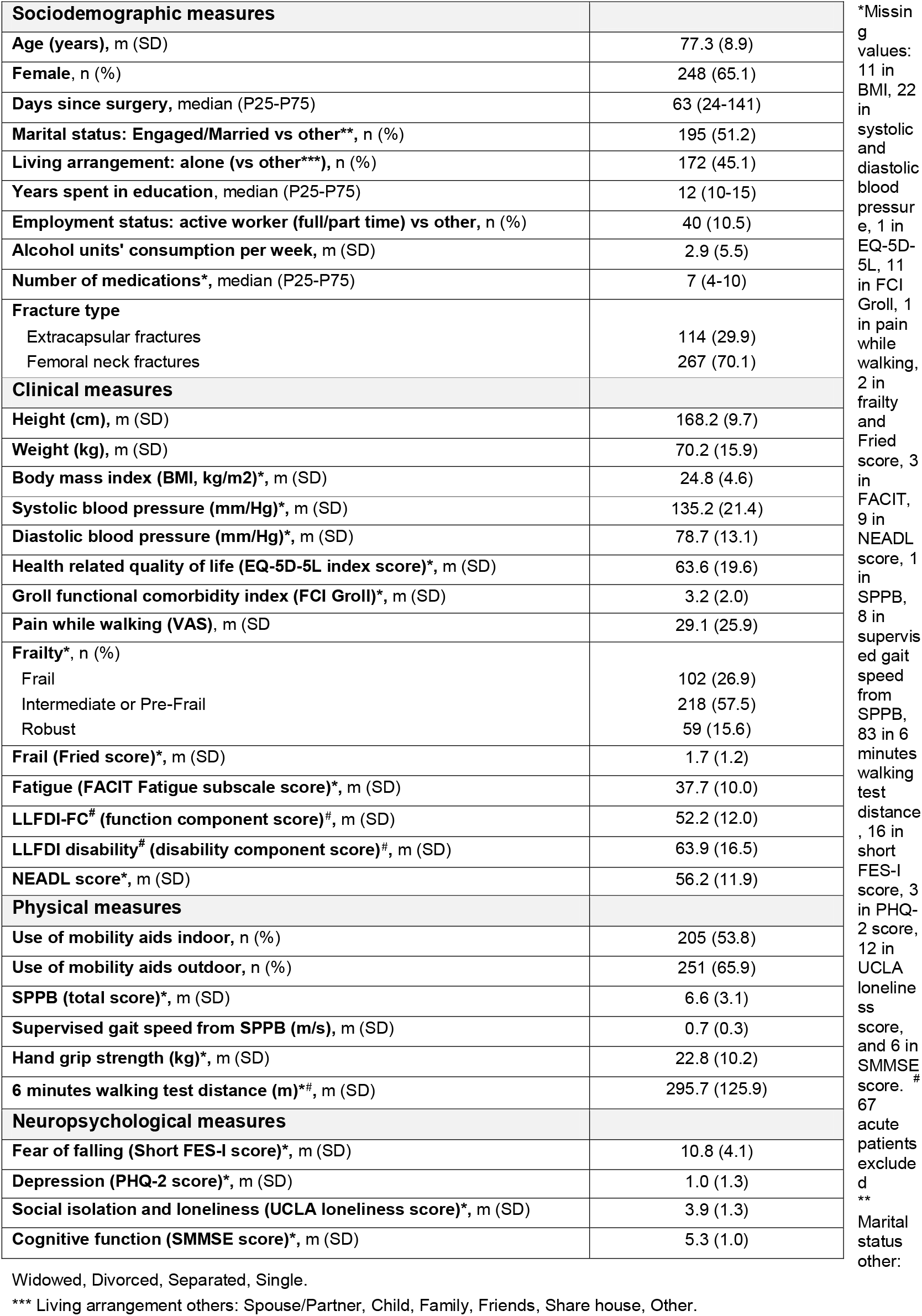
Sociodemographic and clinical characteristics at first visit of included participants with available data at both visits.

| Sociodemographic measures |  |  |
| --- | --- | --- |
| Age (years), m (SD) |  | 77.3 (8.9) |
| Female, n (%) |  | 248 (65.1) |
| Days since surgery, median (P25-P75) |  | 63 (24-141) |
| Marital status: Engaged/Married vs other**, n (%) |  | 195 (51.2) |
| Living arrangement: alone (vs other***), n (%) |  | 172 (45.1) |
| Years spent in education, median (P25-P75) |  | 12 (10-15) |
| Employment status: active worker (full/part time) vs other, n (%) |  | 40 (10.5) |
| Alcohol units' consumption per week, m (SD) |  | 2.9 (5.5) |
| Number of medications*, median (P25-P75) |  | 7 (4-10) |
| Fracture type |  |  |
| Extracapsular fractures |  | 114 (29.9) |
| Femoral neck fractures |  | 267 (70.1) |
| Clinical measures |  |  |
| Height (cm), m (SD) |  | 168.2 (9.7) |
| Weight (kg), m (SD) |  | 70.2 (15.9) |
| Body mass index (BMI, kg/m2)*, m (SD) |  | 24.8 (4.6) |
| Systolic blood pressure (mm/Hg)*, m (SD) |  | 135.2 (21.4) |
| Diastolic blood pressure (mm/Hg)*, m (SD) |  | 78.7 (13.1) |
| Health related quality of life (EQ-5D-5L index score)*, m (SD) |  | 63.6 (19.6) |
| Groll functional comorbidity index (FCI Groll)*, m (SD) |  | 3.2 (2.0) |
| Pain while walking (VAS), m (SD) |  | 29.1 (25.9) |
| Frailty*, n (%) |  |  |
| Frail |  | 102 (26.9) |
| Intermediate or Pre-Frail |  | 218 (57.5) |
| Robust |  | 59 (15.6) |
| Frail (Fried score)*, m (SD) |  | 1.7 (1.2) |
| Fatigue (FACIT Fatigue subscale score)*, m (SD) |  | 37.7 (10.0) |
| LLFDI-FC# (function component score)#, m (SD) |  | 52.2 (12.0) |
| LLFDI disability# (disability component score)#, m (SD) |  | 63.9 (16.5) |
| NEADL score*, m (SD) |  | 56.2 (11.9) |
| Physical measures |  |  |
| Use of mobility aids indoor, n (%) |  | 205 (53.8) |
| Use of mobility aids outdoor, n (%) |  | 251 (65.9) |
| SPPB (total score)*, m (SD) |  | 6.6 (3.1) |
| Supervised gait speed from SPPB (m/s), m (SD) |  | 0.7 (0.3) |
| Hand grip strength (kg)*, m (SD) |  | 22.8 (10.2) |
| 6 minutes walking test distance (m)*#, m (SD) |  | 295.7 (125.9) |
| Neuropsychological measures |  |  |
| Fear of falling (Short FES-I score)*, m (SD) |  | 10.8 (4.1) |
| Depression (PHQ-2 score)*, m (SD) |  | 1.0 (1.3) |
| Social isolation and loneliness (UCLA loneliness score)*, m (SD) |  | 3.9 (1.3) |
| Cognitive function (SMMSE score)*, m (SD) |  | 5.3 (1.0) |

\*Missing values: 11 in BMI, 22 in systolic and diastolic blood pressure, 1 in EQ-5D-5L, 11 in FCI Groll, 1 in pain while walking, 2 in frailty and Fried score, 3 in FACIT, 9 in NEADL score, 1 in SPPB, 8 in supervised gait speed from SPPB, 83 in 6 minutes walking test distance, 16 in short FES-I score, 3 in PHQ-2 score, 12 in UCLA loneliness score, and 6 in SMMSE score. #
67 acute patients excluded
Marital status
Widowed, Divorced, Separated, Single.
\*\*\* Living arrangement others: Spouse/Partner, Child, Family, Friends, Share house, Other.

The DMO values at first and 6-month follow-up visits and the change scores are shown in Table 2.

**Table 2.**
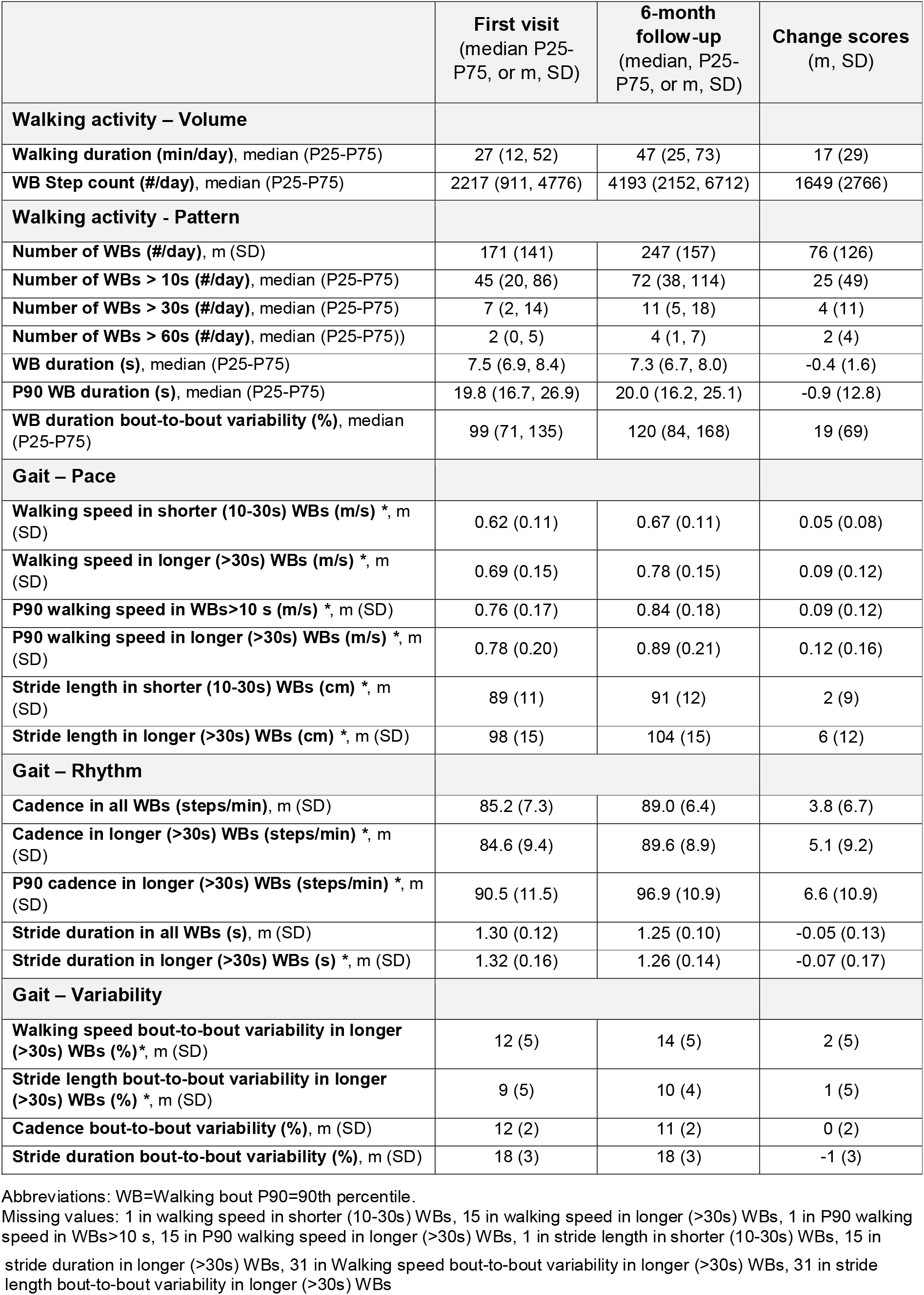
Distribution of DMOs at first visit, at 6-month-follow-up and change scores (n=381)

Table 3 shows the distribution of the anchor variables. Most participants assessed their walking as a little better (28%) or much better (33%) compared to six months earlier (GIC question). This improvement was reflected in the corresponding changes in the three mobility-related outcomes. The remaining 39% of participants perceived their walking as unchanged (13%), a little worse (6.8%), much worse (2.9%), and 17% had a missing value. Overall, the changes in the anchor variables were substantial and above the established MIDs.

**Table 3.** Descriptives of anchor variables at first visit, at 6-month follow-up and change scores (n=381)

|  | First visit | 6-month follow-up | Change scores |
| --- | --- | --- | --- |
| **Global impression of change (GIC) #** |  |  |  |
| Much worse (n, %) |  | 11 (2.9%) |  |
| A little worse (n, %) |  | 26 (6.8%) |  |
| The same (no change) (n, %) |  | 49 (13%) |  |
| A little better (n, %) |  | 105 (28%) |  |
| Much better (n, %) |  | 126 (33%) |  |
| Missing (n, %) |  | 64 (17%) |  |
| **Mobility-related outcome measures** |  |  |  |
| **LLFDI-FC (function component score), m (SD) \*** | 52.1 (12.0) | 57.5 (13.3) | 5.3 (8.5) |
| **SPPB (total score), m (SD) \*** | 6.6 (3.1) | 8.3 (2.9) | 1.8 (2.6) |
| **Supervised gait speed from SPPB (m/s), m (SD) \*** | 0.67 (0.33) | 0.82 (0.31) | 0.15 (0.28) |

Figure 1 provides an overall picture of the course of the four anchors and presents the effect sizes (standardized mean changes) of the three mobility-related outcome measures plotted against the corresponding GIC category. Participants who rated their walking at the 6-month follow-up as little or much better also demonstrated moderate to large positive effect sizes (standardized changes) across the three anchor outcomes. Contrary to expectation, small positive effect sizes were also observed among participants who perceived their walking as little worse or unchanged (except for supervised gait speed). Hence, although improvements were detected in these measures, they were not fully reflected in participants perceived overall mobility.

**Figure 1.**
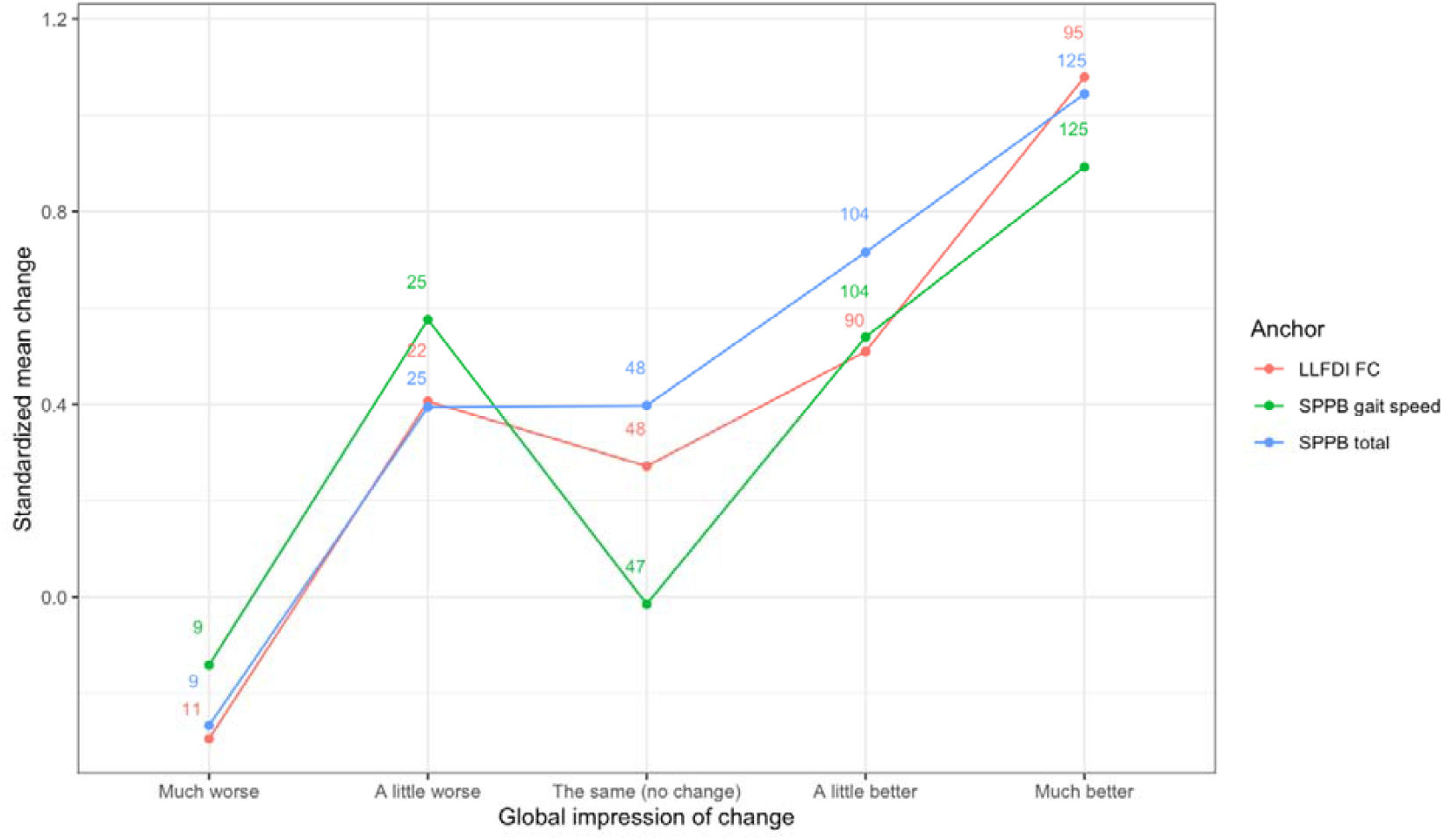
Overall picture of the course of the four anchors: Effect sizes (standardized mean change) of the change of the mobility-related outcome measures by GIC category. Numbers denote the number of measurements per instrument and GIC category.

### Ability to detect changes

Except for the five bout-to-bout variability DMOs and the two WB duration DMOs, the DMOs showed at least moderate positive effects (standardized mean changes >0.5) in the expected direction across the four anchors in those persons who improved in the anchor variables. Seven DMOs exhibited *moderate effects* to detect changes (standardized mean changes 0.5-0.8): Number of WBs >30s and >60s, stride length in shorter (10-30s) WBs, cadence in all WBs, cadence in longer (>30s) WBs, stride duration in all WBs, stride duration in longer (>30s) WBs. Ten DMOs showed *large effects* (standardized mean changes >0.8) to detect changes: Walking duration, WB step count, number of WBs, number of WBs >10s, walking speed in shorter (10-30s) and longer (>30s) WBs, P90 walking speed in WB >10s, P90 walking speed in longer (>30s) WBs, stride length in longer (>30s) WBs, P90 cadence in longer (>30s) WB. In many DMOs slight positive effects were also shown in those persons who did not change in the anchor variables. In some of the DMOs, slight positive effects were shown in persons who worsened in the respective anchors. The detailed results of the DMOs’ changes stratified by the anchors are presented in the Supplementary Material (Tables S3-S6 and Figures S1-S4). Table S7 provides a colour-coded summary of all effect sizes (standardized mean changes) per DMO.

### Minimal Important Difference

Thirty-six of 72 DMO-anchor pairs met the criteria for calculating linear model-based MID estimates (Table S8), and 10 of 96 pairs met the criteria for ROC-based estimates (Table S9). Sufficient anchor-based estimates were available for 12 DMOs, enabling MID triangulation. An overview of all estimates and triangulation results is presented in Table 4. The triangulated estimations of MIDs for walking activity were a walking duration 10 minutes and a WB step count of 1,000 steps per day. MIDs for the category pattern were 50 WBs per day and 15 WBs >10 seconds per day. MIDs for gait pace (i.e., walking speed) were 0.04 m/s in shorter WBs (10-30s), 0.08 m/s in longer WBs >30s, 0.07 m/s in P90 in WBs >10s, 0.08 m/s in P90 in WBs >30s, and stride length of 5 cm in WBs >30s. Lastly, MIDs for gait rhythm were cadence of 4 steps/minute in WBs >30s, cadence of 6 in P90 in WBs >30s, and stride duration of 0.05 seconds in WBs >30s.

**Table 4.**
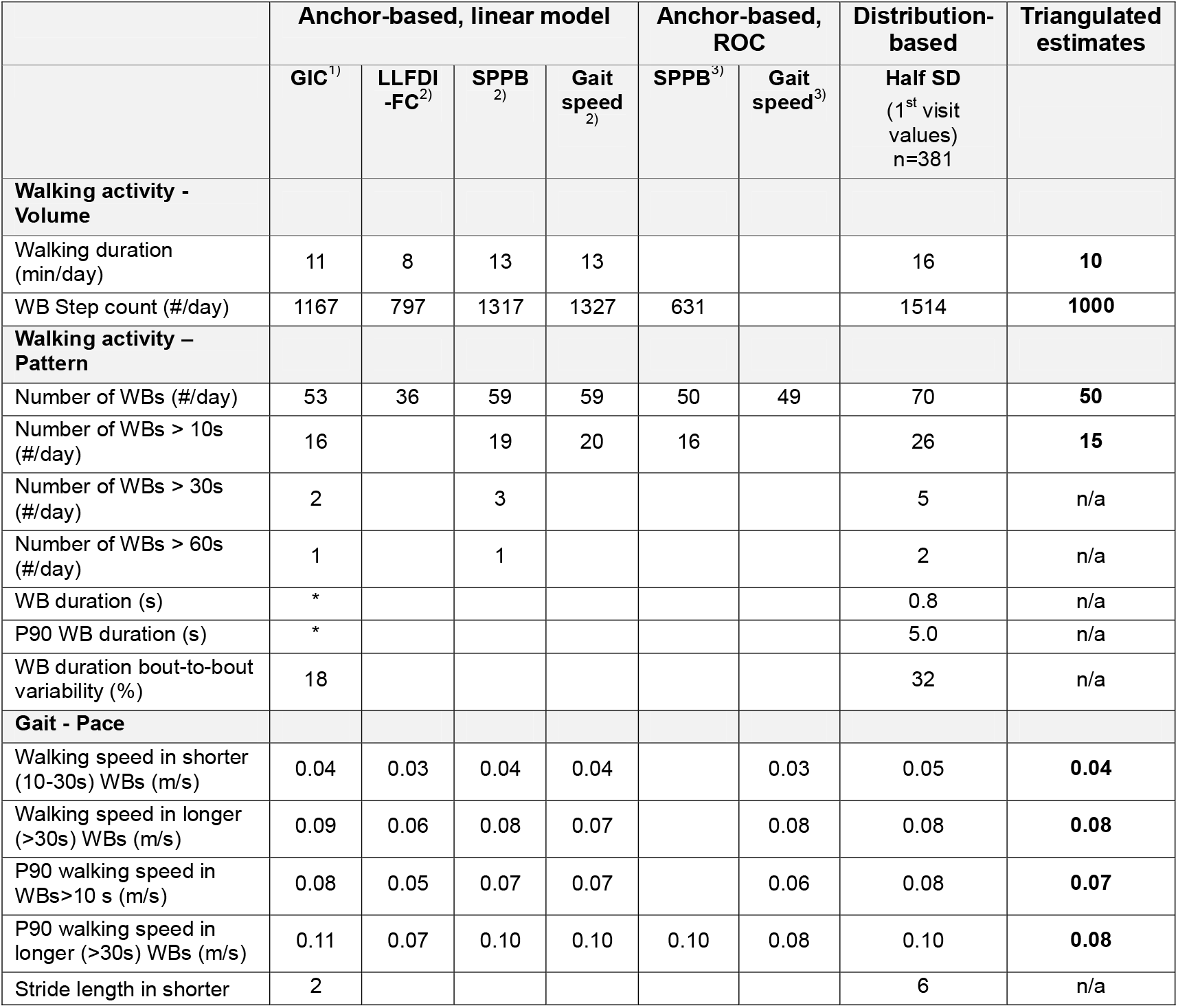
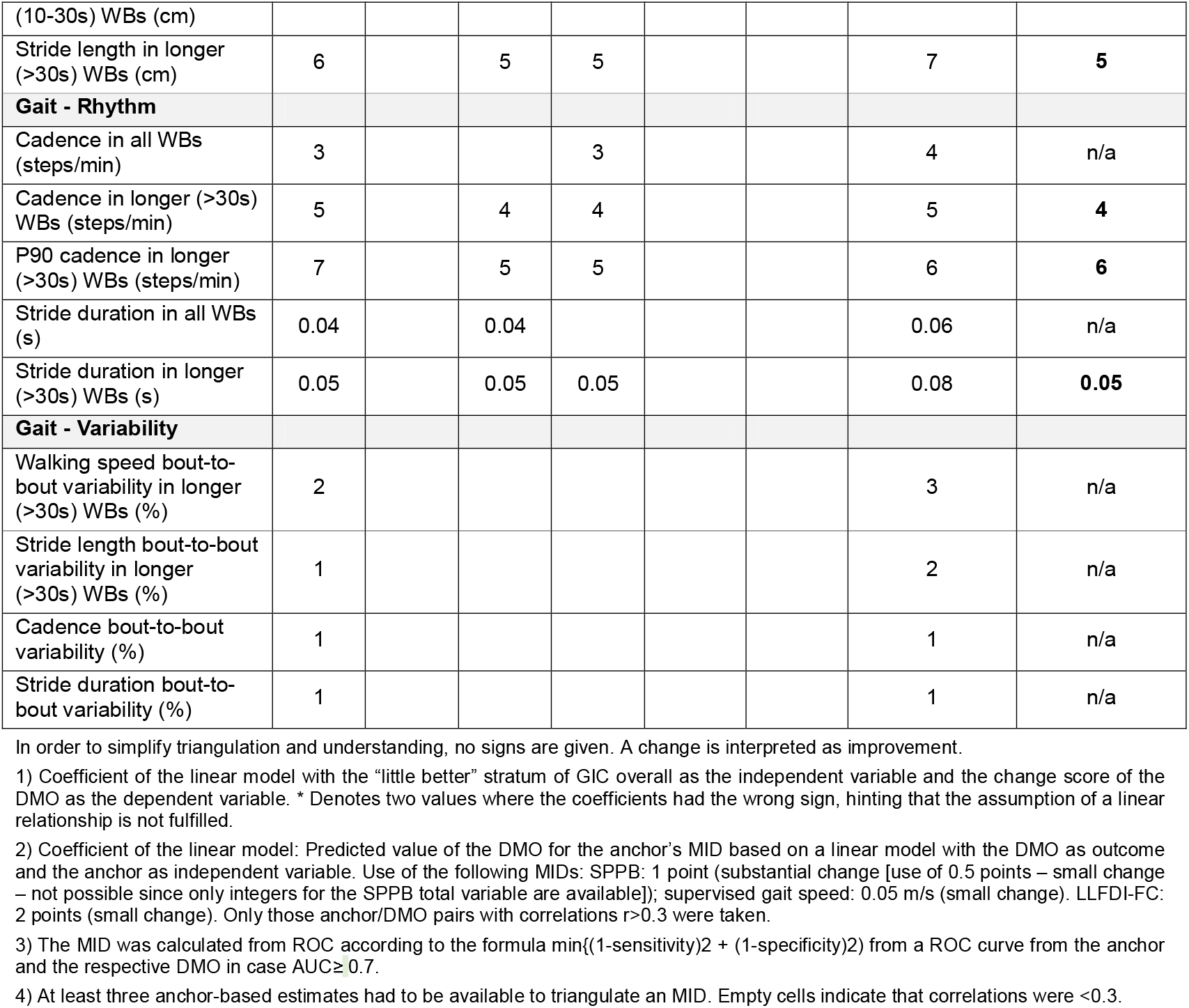
Overview of estimated MID values for the PFF cohort and the triangulated MID estimates.

## Discussion

This work is the first to present results on real-world DMOs’ ability to detect change and the minimal important difference estimates in patients after a PFF. Almost all DMOs detected positive changes in those participants who perceived improvements, with mostly moderate to large effects, indicating that improvement in mobility is substantial during recovery from PFF surgery. However, many DMOs also detected positive changes contrary to expectations in participants who perceived worsening.

### Ability to detect changes

A majority of the DMOs showed a strong ability to detect improvement in mobility, but rarely in detecting decline. In particular, seven of 24 evaluated DMOs showed moderate, and ten DMOs large positive effects across anchors. It is the detection of improvement that is of higher value in this target group, as PFF patients generally can be expected to show improvements in mobility in the first half year after surgery and beyond (34). With a median of 100 days since surgery, most participants can be expected to have been on this upward trajectory as previously shown (34), which is reflected by the high proportion of 60% to 65% of those who reported improvements in all anchors used in this study. Many DMOs detected small positive effects despite some patients reporting subjective worsening according to GIC and LLFDI-FC. This means that even if participants perceived their mobility to be worsening, many DMOs showed improvements. This is not implausible, as they may have had different recovery expectations or a general recovery dissatisfaction, i.e., recovery expectations might affect this relationship. Unfortunately, actions to manage recovery expectations and rehabilitation expectations have not yet been in focus (35). In addition, many patients seem to decline over time in functional aspects that are not covered by the DMOs applied in the present study, such as functional independence in activities of daily living (36). Furthermore, a significant proportion of older adults who experience a hip fracture do not regain their pre-fracture mobility levels (12) – a fact that might entail disappointment and negatively bias perception of their mobility in those who do not reach this level. Apart from mobility, other areas of individual life are affected as well, such as social functioning, cognition, or instrumental task performance (37), all of which are related to individuals’ quality of life and hence may shift patients’ perceived mobility to the negative.

### Minimal Important Difference

Generally, the MID is used to facilitate interpretation and to enable an understanding of treatment results when looking at patient outcome measures (20). So far, only single studies have looked at MIDs in PFF fracture patients. Arcolin et al. (38) were able to establish the MID of the Functional Independence Measure for assessing functional recovery and independence in older adults after PFF. In the present work, we estimated the MID of real-world DMOs. Although these are not yet established for the evaluation of treatment or rehabilitation outcomes, projects such as Mobilise-D (www.mobilise-d.eu) have started to connect DMOs to clinical outcomes for regulatory and clinical endorsement. For twelve DMOs, at least three anchor-based estimates were available following the hierarchy of robustness outlined in the methods section, and MIDs for these twelve DMOs were triangulated. For walking activity, changes in walking duration of 10 minutes and 1,000 steps per day were established as MIDs. These values represent realistic goals for increases in walking for a population with mobility limitations as the current cohort of PFF. The face validity of communication of MIDs was taken into consideration as well, and public awareness is much easier to reach for a value of 1,000 than, e.g., for 950 steps per day. Triangulation of walking pattern parameters identified MIDs of 50 WBs per day and 15 WBs with at least 10 seconds duration. In both parameters, MIDs represent about a third of the overall number of WBs at baseline, about 20% of follow-up values, and are about 2/3 of the actual change between measurements. Hence, the majority of participants actually achieved this MID within 6 months. Positive effects of getting up more frequently have been shown on crucial cardiometabolic factors (39).

The Mobilise-D processing pipeline distinguishes between WBs of >10 s, 10-30 s, and >30 s. MIDs for mean durations and 90^th^ percentiles of these WBs range between 0.04 and 0.08 m/s. MIDs of mean and P90 walking speed in WBs >30 s are identical in this target group. This might be related to longer WBs coming close to their maximum capacity, which is underlined by only a median of 3.7 of these WBs being executed per day overall. Stride length in WBs >30 s had a MID of 5 cm, which is about 6% of the baseline value. As this MID value is smaller than the probable measurement error (19), stride length cannot yet be seen as a useful parameter to determine clinically meaningful changes over time when using a sensor setup as in the present study.

The remaining gait rhythm-based MIDs were cadence changes of 4 steps per minute in >30 s WBs and P90 cadence in >30 s WBs of 6 steps per minute. These values are very close to the actual changes in six months of 5.1 steps per minute in >30 s WBs and 6.6 in P90. This is similar to results for stride duration in >30 s WBs. Again, it must be kept in mind that such “longer” WBs rarely occur in this sample due to their limited mobility.

### Implications for Research and Regulatory Bodies

The present findings come with high practical relevance on several levels. For research, it is key to know the evaluative properties of DMOs. It supports the selection of DMOs with the ability to detect change as endpoints in clinical trials and informs sample size calculations based on the MID as well as interpretation of changes over time and treatment effects. For clinicians, the DMOs provide reassurance that a patient is on an expected positive trajectory or can trigger a visit if a patient is underperforming. Since many DMOs can detect change over time, DMOs may be useful for patients to better interpret their own trajectories and – as a result – to take shared decisions with clinicians. Since mobility is the aspect that is affected the most by a PFF, it is also the most important and meaningful aspect to understand one’s own recovery. On the side of regulatory bodies, the existence of MIDs is an important step for the evaluation of therapy and rehabilitation. The present findings support the use of real-world mobility as a primary endpoint along with PROMs to assess core domains such pain, mood and fatigue.

### Strengths and Limitations

A strength of the current study is that we included multiple professions, i.e. geriatricians, orthopedic surgeons, movement scientists, physiotherapists, psychologists and epidemiologists from different service models in the triangulation process. This ensured that diverse views and broad expertise were taken into account for the establishment of MIDs. Furthermore, the Mobilise-D patient and public advisory group reviewed the GIC question a priori to the implementation into the study, maximizing the patient and patient advocacy input. In contrast to many othe studies, we stated strict a priori criteria and only triangulated MIDs if at least 3 anchor-based estimates were available.

Some potential weaknesses should be highlighted as well. The presented DMOs are all looking at gait and walking which is a core part of mobility but does not represent all positional changes. Further work should therefore include the validation of additional DMOs to measure, e.g. sit-to-stand transfers, sedentary time, and standing time to develop a fully comprehensive model of mobility. The accuracy of spatial parameters such as stride length is not yet optimal in the acute phase and should be further improved (40). Patients were not stratified according to the time since surgery at study inclusion, which varied considerably across the study population and may have influenced the outcomes. However, this approach allowed us to account for the heterogeneity in study populations in practice. In addidion, the recollection of overall walking as assessed by the GIC question may be affected by recall bias, since some patients may not be able to recall the degree to which they have changed (21). However, we triangulated MIDs only when at least two additional estimates based on changes in clinical outcome anchors were available, and our carefully formulated a priori triangulation criteria accounted for potential recall bias. Finally, we cannot exclude the possibility of selection bias, as only 381 of the 506 participants had data available at both time points and were included in the analysis.

### Conclusion and perspectives

This study was successful in establishing MID values for community-dwelling PFF patients without major cognitive impairment. These can provide guidance for research and regulatory bodies. The work underlines the high potential for DMOs to guide the design of studies and the selection of meaningful DMOs. It can also inform sample size calculations for future studies in PFF trials. Next steps should include the testing of responsiveness in controlled trials of (non-)pharmacological interventions.

## Data Availability

The data presented in this paper will be made available under a CC4.0 license in June 2026. Upon release, it can be accessed at: https://zenodo.org/communities/mobilise-d/

## Acknowledgements

This work was supported by the Mobilise-D project that has received funding from the Innovative Medicines Initiative (IMI) 2 Joint Undertaking (JU) under grant agreement number 820820. This JU receives support from the European Union’s Horizon 2020 research and innovation programme and the European Federation of Pharmaceutical Industries and Associations (EFPIA). The content of the current publication reflects the authors’ view, and neither IMI nor the European Union, EFPIA or any Associated Partners are responsible for any use that may be made of the information contained herein. We thank the Mobilise-D consortium for their strong dedication and hard work on this project and our common goal to provide impactful DMOs for clinical populations. We also thank our patients who were willing to take part in the assessments and were as dedicated as we were to our work. SDD and LR were supported by the National Institute for Health and Care Research (NIHR) Newcastle Biomedical Research Centre based at The Newcastle upon Tyne Hospitals NHS Foundation Trust, Newcastle University and the Cumbria, Northumberland and Tyne and Wear (CNTW) NHS Foundation Trust. The research was also supported by NIHR Newcastle Clinical Research Facility (CRF) Infrastructure funding. SDD and LR were also supported by the Innovative Medicines Initiative 2 Joint Undertaking (IMI2 JU) project IDEA-FAST - Grant Agreement 853981. SDD and LR were supported by the UK Research and Innovation (UKRI) Engineering and Physical Sciences Research Council (EPSRC) (Grant Ref: EP/X031012/1 and Grant Ref: EP/X036146/1).

## References

1. Jaeger SU, Wohlrab M, Schoene D, Tremmel R, Chambers M, Leocani L, et al. Mobility endpoints in marketing authorisation of drugs: what gets the European medicines agency moving? Age and ageing. 2022;51(1).

2. Bernabei R, Landi F, Calvani R, Cesari M, Del Signore S, Anker SD, et al. Multicomponent intervention to prevent mobility disability in frail older adults: randomised controlled trial (SPRINTT project). BMJ (Clinical research ed). 2022;377:e068788.

3. Viceconti M, Tome M, Dartee W, Knezevic I, Hernandez Penna S, Mazzà C, et al. On the use of wearable sensors as mobility biomarkers in the marketing authorization of new drugs: A regulatory perspective. Front Med (Lausanne). 2022;9:996903.

4. Rochester L, Mazzà C, Mueller A, Caulfield B, McCarthy M, Becker C, et al. A Roadmap to Inform Development, Validation and Approval of Digital Mobility Outcomes: The Mobilise-D Approach. Digit Biomark. 2020;4(Suppl 1):13–27.

5. Takakusaki K. Functional Neuroanatomy for Posture and Gait Control. J Mov Disord. 2017;10(1):1–17.

6. Fritz S, Lusardi M. White paper: “walking speed: the sixth vital sign”. J Geriatr Phys Ther. 2009;32(2):46–9.

7. Studenski S, Perera S, Patel K, Rosano C, Faulkner K, Inzitari M, et al. Gait speed and survival in older adults. Jama. 2011;305(1):50–8.

8. Perera S, Patel KV, Rosano C, Rubin SM, Satterfield S, Harris T, et al. Gait Speed Predicts Incident Disability: A Pooled Analysis. J Gerontol A Biol Sci Med Sci. 2016;71(1):63–71.

9. Cooper C, Cole ZA, Holroyd CR, Earl SC, Harvey NC, Dennison EM, et al. Secular trends in the incidence of hip and other osteoporotic fractures. Osteoporosis international : a journal established as result of cooperation between the European Foundation for Osteoporosis and the National Osteoporosis Foundation of the USA. 2011;22(5):1277–88.

10. Dargent-Molina P, Favier F, Grandjean H, Baudoin C, Schott AM, Hausherr E, et al. Fall-related factors and risk of hip fracture: the EPIDOS prospective study. Lancet (London, England). 1996;348(9021):145–9.

11. Moerman S, Mathijssen NM, Tuinebreijer WE, Nelissen RG, Vochteloo AJ. Less than one-third of hip fracture patients return to their prefracture level of instrumental activities of daily living in a prospective cohort study of 480 patients. Geriatr Gerontol Int. 2018;18(8):1244–8.

12. Dyer SM, Crotty M, Fairhall N, Magaziner J, Beaupre LA, Cameron ID, et al. A critical review of the long-term disability outcomes following hip fracture. BMC geriatrics. 2016;16:158.

13. Gregori G, Johansson L, Axelsson KF, Jaiswal R, Litsne H, Larsson BAM, et al. The role of different physical function tests for the prediction of fracture risk in older women. J Cachexia Sarcopenia Muscle. 2024;15(4):1511–9.

14. Iosifidis M, Iliopoulos E, Panagiotou A, Apostolidis K, Traios S, Giantsis G. Walking ability before and after a hip fracture in elderly predict greater long-term survivorship. J Orthop Sci. 2016;21(1):48–52.

15. Mikolaizak AS, Rochester L, Maetzler W, Sharrack B, Demeyer H, Mazzà C, et al. Connecting real-world digital mobility assessment to clinical outcomes for regulatory and clinical endorsement-the Mobilise-D study protocol. PloS one. 2022;17(10):e0269615.

16. Kirk C, Küderle A, Micó-Amigo ME, Bonci T, Paraschiv-Ionescu A, Ullrich M, et al. Mobilise-D insights to estimate real-world walking speed in multiple conditions with a wearable device. Sci Rep. 2024;14(1):1754.

17. Polhemus AM, Bergquist R, Bosch de Basea M, Brittain G, Buttery SC, Chynkiamis N, et al. Walking-related digital mobility outcomes as clinical trial endpoint measures: protocol for a scoping review. BMJ Open. 2020;10(7):e038704.

18. Viceconti M, Hernandez Penna S, Dartee W, Mazzà C, Caulfield B, Becker C, et al. Toward a Regulatory Qualification of Real-World Mobility Performance Biomarkers in Parkinson’s Patients Using Digital Mobility Outcomes. Sensors (Basel). 2020;20(20).

19. Micó-Amigo ME, Bonci T, Paraschiv-Ionescu A, Ullrich M, Kirk C, Soltani A, et al. Assessing real-world gait with digital technology? Validation, insights and recommendations from the Mobilise-D consortium. J Neuroeng Rehabil. 2023;20(1):78.

20. Carrasco-Labra A, Devji T, Qasim A, Phillips MR, Wang Y, Johnston BC, et al. Minimal important difference estimates for patient-reported outcomes: A systematic survey. J Clin Epidemiol. 2021;133:61–71.

21. Moosmayer S. Use of the minimal important difference as a criterion for clinical importance-are we off track? JSES Rev Rep Tech. 2023;3(1):56–9.

22. Buekers J, Chernova J, Koch S, Marchena J, Lemos J, Becker C, et al. Digital assessment of real-world walking in people with impaired mobility: How many hours and days are needed? Int J Behav Nutr Phys Act. 2025;22(1):148.

23. Buekers J, Chernova J, Marchena J, Koch S, Lemos J, Becker C, et al. Reliability of real-world walking activity and gait assessment in people with COPD. How many hours and days are needed? European Respiratory Journal. 64(suppl 68):PA791.

24. Koch S, Buekers J, Cobo I, Lemos J, Marchena J, Bonci T, et al. From high-resolution time series to a single, clinically-interpretable value - considerations for the aggregation of real world walking speed assessed by wearable sensors in patients with chronic obstructive pulmonary disease (COPD). European Respiratory Journal. 62(suppl 67):PA1595.

25. Wild D, Grove A, Martin M, Eremenco S, McElroy S, Verjee-Lorenz A, et al. Principles of Good Practice for the Translation and Cultural Adaptation Process for Patient-Reported Outcomes (PRO) Measures: report of the ISPOR Task Force for Translation and Cultural Adaptation. Value in health : the journal of the International Society for Pharmacoeconomics and Outcomes Research. 2005;8(2):94–104.

26. Beauchamp MK, Schmidt CT, Pedersen MM, Bean JF, Jette AM. Psychometric properties of the Late-Life Function and Disability Instrument: a systematic review. BMC geriatrics. 2014;14:12.

27. Beauchamp MK, Ward RE, Jette AM, Bean JF. Meaningful Change Estimates for the Late-Life Function and Disability Instrument in Older Adults. J Gerontol A Biol Sci Med Sci. 2019;74(4):556–9.

28. Perera S, Mody SH, Woodman RC, Studenski SA. Meaningful change and responsiveness in common physical performance measures in older adults. Journal of the American Geriatrics Society. 2006;54(5):743–9.

29. Bohannon RW, Glenney SS. Minimal clinically important difference for change in comfortable gait speed of adults with pathology: a systematic review. J Eval Clin Pract. 2014;20(4):295–300.

30. Cohen J. Statistical Power Analysis for the Behavioral Sciences: Routledge; 1988.

31. Schünemann HJ, Guyatt GH. Commentary--goodbye M(C)ID! Hello MID, where do you come from? Health Serv Res. 2005;40(2):593–7.

32. Schünemann HJ, Puhan M, Goldstein R, Jaeschke R, Guyatt GH. Measurement properties and interpretability of the Chronic respiratory disease questionnaire (CRQ). Copd. 2005;2(1):81–9.

33. Perkins NJ, Schisterman EF. The inconsistency of “optimal” cutpoints obtained using two criteria based on the receiver operating characteristic curve. American journal of epidemiology. 2006;163(7):670–5.

34. Engdal M, Taraldsen K, Jansen CP, Peter RS, Vereijken B, Becker C, et al. Real-world mobility recovery after hip fracture: secondary analyses of digital mobility outcomes from four randomized controlled trials. Age Ageing. 2024;53(10).

35. Krakers SM, Woudsma S, van Dartel D, Vermeer M, Vollenbroek-Hutten MMR, Hegeman JH, et al. Rehabilitation of Frail Older Adults after Hip Fracture Surgery: Predictors for the Length of Geriatric Rehabilitation Stay at a Skilled Nursing Home. J Clin Med. 2024;13(15).

36. Hershkovitz A, Pulatov I, Brill S, Beloosesky Y. Can hip-fractured elderly patients maintain their rehabilitation achievements after 1 year? Disabil Rehabil. 2012;34(4):304–10.

37. Magaziner J, Hawkes W, Hebel JR, Zimmerman SI, Fox KM, Dolan M, et al. Recovery from hip fracture in eight areas of function. J Gerontol A Biol Sci Med Sci. 2000;55(9):M498–507.

38. Arcolin I, Godi M, Giardini M, Guglielmetti S, Bellotti L, Corna S. Minimal clinically important difference of the functional independence measure in older adults with hip fracture. Disabil Rehabil. 2024;46(4):812–9.

39. Carson V, Wong SL, Winkler E, Healy GN, Colley RC, Tremblay MS. Patterns of sedentary time and cardiometabolic risk among Canadian adults. Prev Med. 2014;65:23–7.

40. Berge MA, Paraschiv-Ionescu A, Kirk C, Küderle A, Micó-Amigo E, Becker C, et al. Evaluating the Accuracy and Reliability of Real-World Digital Mobility Outcomes in Older Adults After Hip Fracture: Cross-Sectional Observational Study. JMIR Form Res. 2025;9:e67792.

