## Supplement for "Ability to Detect Changes and Minimal Important Difference of Real-World Digital Mobility Outcomes in Proximal Femoral Fracture Patients"

**Supplementary Material**

***Methods***

***Digital Mobility Outcomes***

**Table S1**. Description of the 24 digital mobility outcomes (DMOs) as presented in Delgado-Ortiz (2025), Eur Respir J. 2025 Jul 24;66(1):2402303. doi: 10.1183/13993003.02303-2024.

| **DMOs** | **Definition** | **Unit** |
| --- | --- | --- |
| **Amount** | | |
| Walking duration | Weekly mean of time spent walking per day | min/day |
| WB Step Count | Weekly mean of the number of steps per day | steps/day |
| **Pattern** | | |
| Number of WBs | Weekly mean of the sum of walking bouts per day | WBs/day |
| Number of WBs > 10s | Weekly mean of the sum of walking bouts per day including bouts longer than 10 seconds | WBs/day |
| Number of WBs > 30s | Weekly mean of the sum of walking bouts per day including bouts longer than 30 seconds | WBs/day |
| Number of WBs > 60s | Weekly mean of the sum of walking bouts per day including bouts longer than 60 seconds | WBs/day |
| WB duration | Weekly mean of the daily mean of walking bout duration | s |
| P90 WB duration | Weekly mean of the daily 90th percentile of walking bout duration | s |
| WB duration bout to bout variability | Weekly mean of the daily bout to bout variability of walking bout duration | % |
| **Gait** | | |
| **Pace** |  |  |
| Walking speed in shorter (10-30s) WBs | Weekly mean of the daily average walking speed, assessed in walking bouts between 10 and 30 seconds | m/s |
| Walking speed in longer (>30s) WBs | Weekly mean of the daily average walking speed, assessed in walking bouts longer than 30 seconds | m/s |
| P90 walking speed in WB > 10 s | Weekly mean of the daily 90th percentile of the walking speed, assessed in walking bouts of more than 10 seconds | m/s |
| P90 walking speed in longer (>30s) WB | Weekly mean of the daily 90th percentile of the walking speed, assessed in walking bouts of longer than 30 seconds | m/s |
| Stride length in shorter (10-30s) WBs | Weekly mean of the daily length of two consecutive steps, assessed during walking bouts between 10 and 30 seconds | cm |
| Stride length in longer (>30s) WBs | Weekly mean of the daily average length of two consecutive steps, assessed during walking bouts longer than 30 seconds | cm |
| **Rhythm** |  |  |
| Cadence in all WB | Weekly mean of the daily average of the steps frequency during a period of time (minutes), calculated in all walking bouts | steps/min |
| Cadence in longer (>30s) WBs | Weekly mean of the daily average of the steps frequency during a period of time (minutes), calculated in walking bouts longer than 30 seconds | steps/min |
| P90 cadence in longer (>30s) WB | Weekly mean of the daily 90th percentile of the steps frequency during a period of time (minutes), calculated in walking bouts longer than 30 seconds | steps/min |
| Stride duration in all WBs | Weekly mean of the daily average of the time elapsed between the initial contacts of two consecutive footfalls of the same foot, assessed in all walking bouts | s |
| Stride duration in longer (>30s) WBs | Weekly mean of the daily average of the time elapsed between the initial contacts of two consecutive footfalls of the same foot, assessed in walking bouts longer than 30 seconds | s |
| **Bout to bout variability** |  |  |
| Walking speed bout to bout variability in longer (>30s) WBs | Weekly mean of the daily bout to bout variability of walking speed, assessed in walking bouts of longer than 30 seconds | % |
| Stride length bout to bout variability in longer (>30s) WBs | Weekly mean of the daily bout to bout variability of stride length, assessed in walking bouts of longer than 30 | % |
| Cadence bout to bout variability | Weekly mean of the daily bout to bout variability of cadence, assessed in all walking bouts | % |
| Stride duration bout to bout variability | Weekly mean of the daily bout to bout variability of stride duration, assessed in all walking bouts | % |

**Results**

***Participant and measurement characteristics***

**Table S2**. Sociodemographic and clinical characteristics of included and excluded participants, at first visit

|  | **Included  (n=381)** | **Excluded (n=125)** | **Total (n=506)** |
| --- | --- | --- | --- |
| **Sociodemographic measures** |  |  |  |
| **Age (years),** m (SD) | 77.3 (8.9) | 78.4 (10.9) | 77.5 (9.4) |
| **Female**, n (%) | 248 (65.1) | 85 (68.0) | 333 (65.8) |
| **Days since surgery,** m (SD) | 100.6 (104.3) | 92.5 (101.9) | 98.6 (103.7) |
| **Marital status: Engaged/Married vs other**,** n (%) | 195 (51.2) | 51 (40.8) | 246 (48.6) |
| **Living arrangement: alone (vs other***),** n (%) | 172 (45.1) | 66 (52.8) | 238 (47.0) |
| **Years spent in education**, median (P25-P75) | 12 (10-15) | 11 (9-14) | 12 (10-15) |
| **Employment status: active worker (full/part time) vs other,** n (%) | 40 (10.5) | 14 (11.2) | 54 (10.7) |
| **Alcohol units' consumption per week,** mean (SD) | 2.9 (5.5) | 2.7 (4.8) | 2.8 (5.3) |
| **Number of medications *,** median (P25-P75) | 7 (4-10) | 8 (4-11) | 7 (4-10) |
| **Fracture type**  Extracapsular fractures  Femoral neck fractures | 114 (29.9)  267 (70.1) | 43 (34.4)  82 (65.6) | 157 (31.0)  349 (69.0) |
| **Clinical measures** |  |  |  |
| **Height (cm),** mean (SD) | 168.2 (9.7) | 166.0 (9.9) | 167.7 (9.8) |
| **Weight (kg),** mean (SD) | 70.2 (15.9) | 66.4 (13.9) | 69.3 (15.5) |
| **Body mass index (BMI, kg/m2)*,** m (SD) | 24.8 (4.6) | 24.0 (3.9) | 24.6 (4.5) |
| **Systolic blood pressure (mm/hg) *,** m (SD) | 135.2 (21.4) | 138.6 (23.7) | 136.0 (22.0) |
| **Diastolic blood pressure (mm/hg) *,** m (SD) | 78.7 (13.1) | 78.0 (13.6) | 78.5 (13.2) |
| **Health related quality of life (EQ-5D-5L index score) *,** m (SD) | 63.6 (19.6) | 58.2 (21.8) | 62.3 (20.3) |
| **Groll functional comorbidity index (FCI Groll) *,** m (SD) | 3.2 (2.0) | 3.5 (2.3) | 3.3 (2.1) |
| **Pain while walking (VAS)**, m (SD) | 29.1 (25.9) | 34.3 (26.0) | 30.4 (25.9) |
| **Frailty***, n (%)    Frail    Intermediate or Pre-Frail    Robust | 102 (26.9)  218 (57.5)  59 (15.6) | 42 (34.4)  67 (54.9)  13 (10.7) | 144 (28.7)  285 (56.9)  72 (14.4) |
| **Frail (Fried score)*,** m (SD) | 1.7 (1.2) | 2.1 (1.3) | 1.8 (1.2) |
| **Fatigue (FACIT Fatigue subscale score) *,** m (SD) | 37.7 (10.0) | 36.1 (10.9) | 37.3 (10.2) |
| **LLFDI-FC^#^ (function component score)** ^#^**,** m (SD) | 52.2 (12.0) | 49.0 (12.9) | 51.5 (12.3) |
| **LLFDI disability^#^ (disability component score)** ^#^**,** m (SD) | 63.9 (16.5) | 58.5 (15.6) | 62.7 (16.5) |
| **NEADL score *,** m (SD) | 56.2 (11.9) | 52.5 (14.1) | 55.3 (12.5) |
| **Physical mearures** |  |  |  |
| **Use of mobility aids indoor,** n (%) | 205 (53.8) | 73 (58.4) | 278 (54.9) |
| **Use of mobility aids outdoor,** n (%) | 251 (65.9) | 92 (73.6) | 343 (67.8) |
| **SPPB (total score) *,** m (SD) | 6.6 (3.1) | 5.1 (2.9) | 6.2 (3.1) |
| **Supervised gait speed from SPPB (m/s),** m (SD) | 0.7 (0.3) | 0.6 (0.4) | 0.6 (0.3) |
| **Hand grip strength (kg) *,** m (SD) | 22.8 (10.2) | 19.8 (9.4) | 22.1 (10.1) |
| **6 minutes walking test distance (m) ***^#^**,** m (SD) | 295.7 (125.9) | 238.6 (116.0) | 283.4 (125.9) |
| **Neuropsychological measures** |  |  |  |
| **Fear of falling (Short FES-I score) *,** m (SD) | 10.8 (4.1) | 11.9 (5.0) | 11.1 (4.3) |
| **Depression (PHQ-2 score) *,** m (SD) | 1.0 (1.3) | 1.3 (1.5) | 1.1 (1.4) |
| **Social isolation and loneliness (UCLA loneliness score) *,** m (SD) | 3.9 (1.3) | 4.0 (1.4) | 3.9 (1.4) |
| **Cognitive function (SMMSE score) *,** m (SD) | 5.3 (1.0) | 5.0 (1.2) | 5.2 (1.1) |

*Missing values: 3 in Days since surgery, 20 in BMI, 33 in systolic and diastolic blood pressure, 9 in EQ-5D-5L, 20 in FCI Groll, 7 in pain while walking, 5 in frailty and Fried score, 11 in FACIT, 17 in NEADL score, 3 in SPPB, 11 in supervised gait speed from SPPB, 126 in 6 minutes walking test distance, 25 in short FES-I score, 11 in PHQ-2 score, 21 in UCLA loneliness score, and 10 in SMMSE score. ^#^ 102 acute patients excluded
** Marital status other: Widowed, Divorced, Separated, Single.

*** Living arrangement others: Spouse/Partner, Child, Family, Friends, Share house, Other.

***Ability to detect changes***

In the following, the detailed results of the DMOs’ ability to detect changes are presented, stratified by the categories of the GIC (Table S3, Figure S1), LLFDI function component (Table S4, Figure S2), SPPB total (Table S5, Figure S3) and supervised gait speed (Table S6, Figure S4).

**Table S3.** Change in DMOs from first visit to 6-month follow-up, stratified by Global impression of change scale (GIC) categories

|  | **Global impression of change** | **Mean change** | **SD change** | **Effect size** | **n** |
| --- | --- | --- | --- | --- | --- |
| **Walking activity - Volume** | | | | | |
| **Walking duration (min/day)** | | | | | |
|  | Much worse | -2 | 15 | -0.17 | 11 |
|  | A little worse | 15 | 19 | 0.77 | 26 |
|  | The same (no change) | 7 | 19 | 0.38 | 49 |
|  | A little better | 11 | 24 | 0.47 | 105 |
|  | Much better | 30 | 33 | 0.90 | 126 |
| **WB Step Count (steps/day)** | | | | | |
|  | Much worse | -414 | 1530 | -0.27 | 11 |
|  | A little worse | 1387 | 1772 | 0.78 | 26 |
|  | The same (no change) | 715 | 1774 | 0.40 | 49 |
|  | A little better | 1167 | 2324 | 0.50 | 105 |
|  | Much better | 2881 | 3177 | 0.91 | 126 |
| **Walking activity - Pattern** | | | | | |
| **Number of WBs (#/day)** | | | | | |
|  | Much worse | -32 | 96 | -0.34 | 11 |
|  | A little worse | 83 | 101 | 0.82 | 26 |
|  | The same (no change) | 33 | 79 | 0.41 | 49 |
|  | A little better | 53 | 107 | 0.50 | 105 |
|  | Much better | 125 | 138 | 0.90 | 126 |
| **Number of WBs > 10s (#/day)** | | | | | |
|  | Much worse | 6 | 24 | -0.23 | 11 |
|  | A little worse | 24 | 34 | 0.69 | 26 |
|  | The same (no change) | 14 | 39 | 0.34 | 49 |
|  | A little better | 16 | 46 | 0.34 | 105 |
|  | Much better | 46 | 53 | 0.86 | 126 |
| **Number of WBs > 30s (#/day)** | | | | | |
|  | Much worse | 1 | 5 | 0.16 | 11 |
|  | A little worse | 1 | 7 | 0.19 | 26 |
|  | The same (no change) | 2 | 9 | 0.20 | 49 |
|  | A little better | 2 | 10 | 0.22 | 105 |
|  | Much better | 8 | 13 | 0.64 | 126 |
| **Number of WBs > 60s (#/day)** | | | | | |
|  | Much worse | 1 | 2 | 0.65 | 11 |
|  | A little worse | 1 | 3 | 0.27 | 26 |
|  | The same (no change) | 0 | 4 | 0.10 | 49 |
|  | A little better | 1 | 4 | 0.26 | 105 |
|  | Much better | 3 | 5 | 0.60 | 126 |
| **WB duration (s)** | | | | | |
|  | Much worse | -0.5 | 4.0 | -0.13 | 11 |
|  | A little worse | -0.3 | 1.0 | -0.32 | 26 |
|  | The same (no change) | -0.3 | 1.4 | -0.25 | 49 |
|  | A little better | -0.4 | 1.5 | -0.23 | 105 |
|  | Much better | -0.3 | 1.4 | -0.18 | 126 |
| **P90 WB duration (s)** | | | | | |
|  | Much worse | -0.9 | 10.1 | -0.08 | 11 |
|  | A little worse | -2.6 | 9.4 | -0.27 | 26 |
|  | The same (no change) | -1.3 | 15.5 | -0.08 | 49 |
|  | A little better | -0.9 | 11.0 | -0.09 | 105 |
|  | Much better | 1.27 | 14.1 | 0.09 | 126 |
| **WB duration bout to bout variability** | | | | | |
|  | Much worse | 6 | 34 | 0.18 | 11 |
|  | A little worse | 13 | 38 | 0.35 | 26 |
|  | The same (no change) | 5 | 63 | 0.07 | 49 |
|  | A little better | 17.85 | 61 | 0.29 | 105 |
|  | Much better | 30.09 | 3 | 0.36 | 126 |
| **Gait – Pace** | | | | | |
| **Walking speed in shorter (10-30s) WBs (m/s)** | | | | | |
|  | Much worse | -0.04 | 0.12 | -0.36 | 11 |
|  | A little worse | 0.05 | 0.07 | 0.69 | 25 |
|  | The same (no change) | 0.01 | 0.05 | 0.21 | 49 |
|  | A little better | 0.04 | 0.08 | 0.56 | 105 |
|  | Much better | 0.08 | 0.08 | 1.01 | 126 |
| **Walking speed in longer (>30s) WBs (m/s)** | | | | | |
|  | Much worse | -0.05 | 0.12 | -0.45 | 9 |
|  | A little worse | 0.06 | 0.12 | 0.51 | 23 |
|  | The same (no change) | 0.04 | 0.09 | 0.47 | 45 |
|  | A little better | 0.08 | 0.11 | 0.76 | 100 |
|  | Much better | 0.13 | 0.12 | 1.05 | 117 |
| **P90 walking speed in WBs>10 s (m/s)** | | | | | |
|  | Much worse | -0.04 | 0.15 | -0.27 | 11 |
|  | A little worse | 0.06 | 0.10 | 0.63 | 25 |
|  | The same (no change) | 0.03 | 0.07 | 0.36 | 49 |
|  | A little better | 0.08 | 0.11 | 0.68 | 105 |
|  | Much better | 0.14 | 0.13 | 1.04 | 126 |
| **P90 walking speed in longer (>30s) WBs (m/s)** | | | | | |
|  | Much worse | -0.06 | 0.15 | -0.38 | 9 |
|  | A little worse | 0.08 | 0.16 | 0.51 | 23 |
|  | The same (no change) | 0.06 | 0.11 | 0.53 | 45 |
|  | A little better | 0.11 | 0.15 | 0.75 | 100 |
|  | Much better | 0.17 | 0.16 | 1.05 | 117 |
| **Stride length in shorter (10-30s) WBs (cm)** | | | | | |
|  | Much worse | -5 | 20 | -0.26 | 11 |
|  | A little worse | 3 | 6 | 0.50 | 25 |
|  | The same (no change) | -1 | 8 | -0.16 | 49 |
|  | A little better | 2 | 8 | 0.25 | 105 |
|  | Much better | 5 | 8 | 0.66 | 126 |
| **Stride length in longer (>30s) WBs (cm)** | | | | | |
|  | Much worse | -6 | 20 | -0.32 | 9 |
|  | A little worse | 3 | 11 | 0.26 | 23 |
|  | The same (no change) | 2 | 10 | 0.21 | 45 |
|  | A little better | 6 | 12 | 0.49 | 100 |
|  | Much better | 10 | 12 | 0.85 | 117 |
| **Gait – Rhythm** | | | | | |
| **Cadence in all WBs (steps/min)** | | | | | |
|  | Much worse | -1.7 | 6.3 | -0.27 | 11 |
|  | A little worse | 2.9 | 5.7 | 0.51 | 26 |
|  | The same (no change) | 2.4 | 4.7 | 0.51 | 49 |
|  | A little better | 3.2 | 5.9 | 0.55 | 105 |
|  | Much better | 4.8 | 7.1 | 0.67 | 126 |
| **Cadence in longer (>30s) WBs (steps/min)** | | | | | |
|  | Much worse | -2.7 | 10.3 | -0.27 | 9 |
|  | A little worse | 4.2 | 7.9 | 0.53 | 23 |
|  | The same (no change) | 3.0 | 5.9 | 0.52 | 45 |
|  | A little better | 5.1 | 8.5 | 0.60 | 100 |
|  | Much better | 6.5 | 9.0 | 0.73 | 117 |
| **P90 cadence in longer (>30s) WBs (steps/min)** | | | | | |
|  | Much worse | -4.1 | 12.6 | -0.32 | 9 |
|  | A little worse | 4.8 | 9.3 | 0.51 | 23 |
|  | The same (no change) | 3.8 | 7.0 | 0.54 | 45 |
|  | A little better | 6.5 | 10.1 | 0.65 | 100 |
|  | Much better | 8.9 | 11.0 | 0.80 | 117 |
| **Stride duration in all WBs (s)** | | | | | |
|  | Much worse | 0.05 | 0.18 | 0.26 | 11 |
|  | A little worse | -0.04 | 0.11 | -0.38 | 26 |
|  | The same (no change) | -0.05 | 0.11 | -0.45 | 49 |
|  | A little better | -0.04 | 0.11 | -0.34 | 105 |
|  | Much better | -0.05 | 0.14 | -0.38 | 126 |
| **Stride duration in longer (>30s) WBs (s)** | | | | | |
|  | Much worse | 0.03 | 0.25 | 0.12 | 9 |
|  | A little worse | -0.08 | 0.14 | -0.54 | 23 |
|  | The same (no change) | -0.06 | 0.14 | -0.43 | 45 |
|  | A little better | -0.05 | 0.16 | -0.34 | 100 |
|  | Much better | -0.08 | 0.17 | -0.49 | 117 |
| **Gait – Variability** | | | | | |
| **Walking speed bout to bout variability in longer (>30s) WBs (%)** | | | | | |
|  | Much worse | 0 | 7 | 0.07 | 8 |
|  | A little worse | 1 | 4 | 0.13 | 22 |
|  | The same (no change) | 1 | 4 | 0.33 | 43 |
|  | A little better | 2 | 6 | 0.26 | 89 |
|  | Much better | 2 | 5 | 0.37 | 115 |
| **Stride length bout to bout variability in longer (>30s) WBs (%)** | | | | | |
|  | Much worse | 3 | 6 | 0.54 | 8 |
|  | A little worse | 0 | 3 | 0.03 | 22 |
|  | The same (no change) | 0 | 4 | 0.01 | 43 |
|  | A little better | 1 | 5 | 0.28 | 89 |
|  | Much better | 1 | 5 | 0.19 | 115 |
| **Cadence bout to bout variability (%)** | | | | | |
|  | Much worse | 0 | 4 | 0.13 | 11 |
|  | A little worse | -1 | 2 | -0.33 | 26 |
|  | The same (no change) | 0 | 2 | 0.06 | 49 |
|  | A little better | -1 | 2 | -0.26 | 105 |
|  | Much better | 0 | 2 | -0.20 | 126 |
| **Stride duration bout to bout variability (%)** | | | | | |
|  | Much worse | 0 | 4 | 0.04 | 11 |
|  | A little worse | -1 | 3 | -0.29 | 26 |
|  | The same (no change) | 1 | 3 | 0.20 | 49 |
|  | A little better | -1 | 3 | -0.17 | 105 |
|  | Much better | -1 | 3 | -0.41 | 126 |

Colour coding: Green shading shows effect sizes higher than |0.2| and detected changes in the expected direction (light green small effects: |≥0.2 to <0.5|, medium green moderate effects: |≥0.5 to <0.8|, dark green large effects: |≥0.8|). Grey shading shows effect sizes higher than |0.2| and detected changes against expectations. Blue shading shows effect sizes higher than |0.2| in the five bout-to-bout variability DMOs for which we did not have an expectation (with one exception all <0.5). No shading (white) means no effect (effect sizes <0.2).

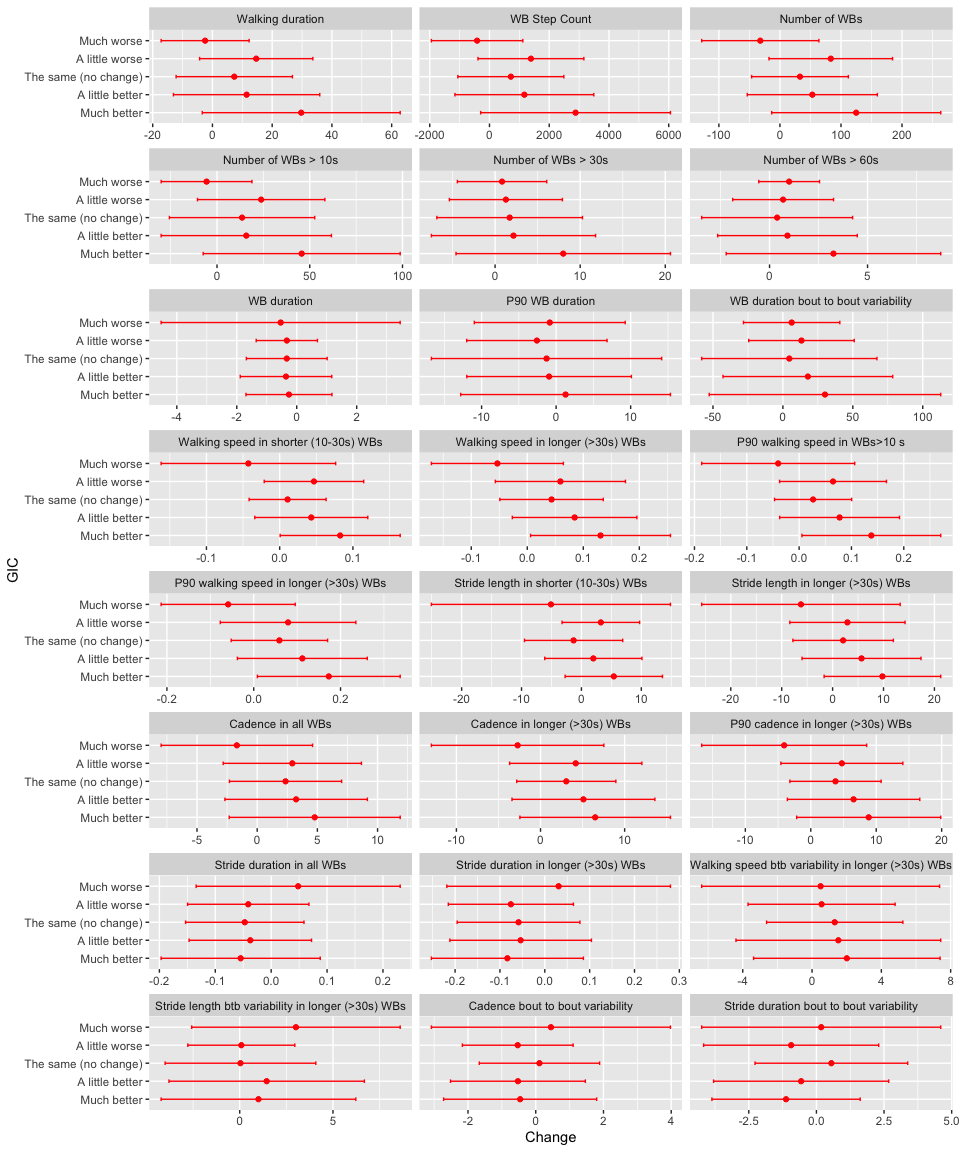

**Figure S1**. Plot of mean changes and standard deviations from first visit to 6-months follow-up, stratified by GIC categories. Btb=Bout to bout

**Table S4**. Change in DMOs from first visit to 6-month follow-up, stratified by LLFDI function component categories

|  | **LLFDI function component ^1)^** | **Mean change** | **SD change** | **Effect size** | **n** |
| --- | --- | --- | --- | --- | --- |
| **Walking activity - Volume** | | | | | |
| **Walking duration (min/day)** | | | | | |
|  | Much worse | 3 | 19 | 0.14 | 28 |
|  | Little worse | 1 | 13 | 0.10 | 24 |
|  | No | 6 | 20 | 0.29 | 62 |
|  | Little better | 3 | 16 | 0.19 | 56 |
|  | Much better | 22 | 31 | 0.69 | 144 |
| **WB Step Count (steps/day)** | | | | | |
|  | Much worse | 313 | 1789 | 0.18 | 28 |
|  | Little worse | 61 | 1235 | 0.05 | 24 |
|  | No | 541 | 1857 | 0.29 | 62 |
|  | Little better | 360 | 1446 | 0.25 | 56 |
|  | Much better | 2227 | 3030 | 0.74 | 144 |
| **Walking activity - Pattern** | | | | | |
| **Number of WBs (#/day)** | | | | | |
|  | Much worse | 1 | 72 | 0.02 | 28 |
|  | Little worse | 2 | 76 | 0.02 | 24 |
|  | No | 32 | 62 | 0.51 | 62 |
|  | Little better | 26 | 81 | 0.33 | 56 |
|  | Much better | 90 | 132 | 0.69 | 144 |
| **Number of WBs > 10s (#/day)** | | | | | |
|  | Much worse | 1 | 32 | 0.04 | 28 |
|  | Little worse | 1 | 23 | 0.06 | 24 |
|  | No | 11 | 28 | 0.39 | 62 |
|  | Little better | 4 | 32 | 0.14 | 56 |
|  | Much better | 32 | 55 | 0.57 | 144 |
| **Number of WBs > 30s (#/day)** | | | | | |
|  | Much worse | 0 | 8 | 0.04 | 28 |
|  | Little worse | 0 | 5 | -0.02 | 24 |
|  | No | 1 | 7 | 0.14 | 62 |
|  | Little better | 0 | 7 | -0.07 | 56 |
|  | Much better | 5 | 13 | 0.41 | 144 |
| **Number of WBs > 60s (#/day)** | | | | | |
|  | Much worse | 0 | 3 | 0.10 | 28 |
|  | Little worse | 0 | 3 | 0.06 | 24 |
|  | No | 0 | 3 | 0.08 | 62 |
|  | Little better | 0 | 3 | -0.09 | 56 |
|  | Much better | 2 | 5 | 0.45 | 144 |
| **WB duration (s)** | | | | | |
|  | Much worse | -0.3 | 2.5 | -0.12 | 28 |
|  | Little worse | -0.5 | 1.8 | -0.25 | 24 |
|  | No | -0.3 | 1.3 | -0.24 | 62 |
|  | Little better | -0.7 | 1.6 | -0.45 | 56 |
|  | Much better | -0.3 | 1.4 | -0.20 | 144 |
| **P90 WB duration (s)** | | | | | |
|  | Much worse | 0.1 | 11.5 | 0.01 | 28 |
|  | Little worse | -0.6 | 14.6 | -0.04 | 24 |
|  | No | -1.8 | 14.1 | -0.13 | 62 |
|  | Little better | -2.7 | 11.7 | -0.23 | 56 |
|  | Much better | -1.2 | 12.4 | -0.10 | 144 |
| **WB duration bout to bout variability (%)** | | | | | |
|  | Much worse | 16 | 52 | 0.31 | 28 |
|  | Little worse | 6 | 79 | 0.08 | 24 |
|  | No | -4 | 67 | -0.06 | 62 |
|  | Little better | -2 | 37 | -0.05 | 56 |
|  | Much better | 22 | 78 | 0.29 | 144 |
| **Gait – Pace** | | | | | |
| **Walking speed in shorter (10-30s) WBs (m/s)** | | | | | |
|  | Much worse | 0.01 | 0.09 | 0.13 | 28 |
|  | Little worse | 0.00 | 0.05 | 0.09 | 24 |
|  | No | 0.02 | 0.05 | 0.34 | 62 |
|  | Little better | 0.02 | 0.06 | 0.40 | 56 |
|  | Much better | 0.06 | 0.07 | 0.79 | 143 |
| **Walking speed in longer (>30s) WBs (m/s)** | | | | | |
|  | Much worse | 0.04 | 0.10 | 0.40 | 26 |
|  | Little worse | 0.02 | 0.11 | 0.21 | 24 |
|  | No | 0.04 | 0.08 | 0.52 | 57 |
|  | Little better | 0.05 | 0.11 | 0.47 | 51 |
|  | Much better | 0.10 | 0.12 | 0.90 | 141 |
| **P90 walking speed in WBs>10 s (m/s)** | | | | | |
|  | Much worse | 0.02 | 0.11 | 0.19 | 28 |
|  | Little worse | 0.01 | 0.09 | 0.13 | 24 |
|  | No | 0.03 | 0.07 | 0.43 | 62 |
|  | Little better | 0.03 | 0.09 | 0.33 | 56 |
|  | Much better | 0.11 | 0.11 | 0.94 | 143 |
| **P90 walking speed in longer (>30s) WBs (m/s)** | | | | | |
|  | Much worse | 0.05 | 0.11 | 0.43 | 26 |
|  | Little worse | 0.03 | 0.14 | 0.21 | 24 |
|  | No | 0.06 | 0.10 | 0.56 | 57 |
|  | Little better | 0.07 | 0.14 | 0.47 | 51 |
|  | Much better | 0.13 | 0.15 | 0.89 | 141 |
| **Stride length in shorter (10-30s) WBs (cm)** | | | | | |
|  | Much worse | -1 | 13 | -0.04 | 28 |
|  | Little worse | -1 | 5 | -0.18 | 24 |
|  | No | 0 | 8 | -0.01 | 62 |
|  | Little better | 1 | 7 | 0.09 | 56 |
|  | Much better | 3 | 7 | 0.39 | 143 |
| **Stride length in longer (>30s) WBs (cm)** | | | | | |
|  | Much worse | 2 | 14 | 0.14 | 26 |
|  | Little worse | 2 | 11 | 0.20 | 24 |
|  | No | 2 | 11 | 0.18 | 57 |
|  | Little better | 4 | 13 | 0.32 | 51 |
|  | Much better | 6 | 10 | 0.64 | 141 |
| **Gait – Rhythm** | | | | | |
| **Cadence in all WBs (steps/min)** | | | | | |
|  | Much worse | 1.3 | 4.3 | 0.31 | 28 |
|  | Little worse | 0.9 | 6.1 | 0.16 | 24 |
|  | No | 2.1 | 4.9 | 0.43 | 62 |
|  | Little better | 2.7 | 4.6 | 0.59 | 56 |
|  | Much better | 4.2 | 6.9 | 0.61 | 144 |
| **Cadence in longer (>30s) WBs (steps/min)** | | | | | |
|  | Much worse | 2.3 | 6.0 | 0.39 | 26 |
|  | Little worse | 0.9 | 6.4 | 0.14 | 24 |
|  | No | 2.9 | 8.3 | 0.35 | 57 |
|  | Little better | 2.9 | 7.4 | 0.39 | 51 |
|  | Much better | 6.2 | 9.6 | 0.65 | 141 |
| **P90 cadence in longer (>30s) WBs (steps/min)** | | | | | |
|  | Much worse | 2.2 | 6.8 | 0.32 | 26 |
|  | Little worse | 0.4 | 6.1 | 0.07 | 24 |
|  | No | 3.7 | 9.6 | 0.38 | 57 |
|  | Little better | 3.4 | 8.5 | 0.41 | 51 |
|  | Much better | 7.6 | 11.0 | 0.69 | 141 |
| **Stride duration in all WBs (s)** | | | | | |
|  | Much worse | -0.01 | 0.11 | -0.07 | 28 |
|  | Little worse | 0.02 | 0.15 | 0.14 | 24 |
|  | No | -0.03 | 0.12 | -0.23 | 62 |
|  | Little better | -0.03 | 0.09 | -0.39 | 56 |
|  | Much better | -0.05 | 0.10 | -0.45 | 144 |
| **Stride duration in longer (>30s) WBs (s)** | | | | | |
|  | Much worse | -0.03 | 0.09 | -0.30 | 26 |
|  | Little worse | 0.04 | 0.20 | 0.22 | 24 |
|  | No | -0.03 | 0.19 | -0.15 | 57 |
|  | Little better | -0.05 | 0.13 | -0.43 | 51 |
|  | Much better | -0.08 | 0.14 | -0.55 | 141 |
| **Gait – Variability** | | | | | |
| **Walking speed bout to bout variability in longer (>30s) WBs (%)** | | | | | |
|  | Much worse | 0 | 4 | 0.07 | 25 |
|  | Little worse | 1 | 5 | 0.21 | 24 |
|  | No | 1 | 6 | 0.21 | 54 |
|  | Little better | 1 | 5 | 0.22 | 45 |
|  | Much better | 1 | 5 | 0.28 | 137 |
| **Stride length bout to bout variability in longer (>30s) WBs (%)** | | | | | |
|  | Much worse | 1 | 4 | 0.22 | 25 |
|  | Little worse | 1 | 5 | 0.30 | 24 |
|  | No | 1 | 5 | 0.12 | 54 |
|  | Little better | 1 | 5 | 0.15 | 45 |
|  | Much better | 1 | 4 | 0.20 | 137 |
| **Cadence bout to bout variability (%)** | | | | | |
|  | Much worse | 0 | 2 | 0.06 | 28 |
|  | Little worse | -1 | 2 | -0.41 | 24 |
|  | No | 0 | 1 | 0.21 | 62 |
|  | Little better | 0 | 2 | -0.20 | 56 |
|  | Much better | 0 | 2 | -0.21 | 144 |
| **Stride duration bout to bout variability (%)** | | | | | |
|  | Much worse | 0 | 3 | 0.04 | 28 |
|  | Little worse | 0 | 3 | -0.11 | 24 |
|  | No | 0 | 3 | 0.02 | 62 |
|  | Little better | 0 | 3 | -0.08 | 56 |
|  | Much better | -1 | 3 | -0.38 | 144 |

Changers are defined as change in LLFDI function component according to the established MID, where a change of at least 2 denotes a small change (little worse, little better) and of at least 5 as substantial change (much worse, much better).

1) Subgroup without acute patients (n=67 excluded).

Colour coding: Green shading shows effect sizes higher than |0.2| and detected changes in the expected direction (light green small effects: |≥0.2 to <0.5|, medium green moderate effects: |≥0.5 to <0.8|, dark green large effects: |≥0.8|). Grey shading shows effect sizes higher than |0.2| and detected changes against expectations. Blue shading shows effect sizes higher than |0.2| in the five bout-to-bout variability DMOs for which we did not have an expectation (with one exception all <0.5). No shading (white) means no effect (effect sizes <0.2).

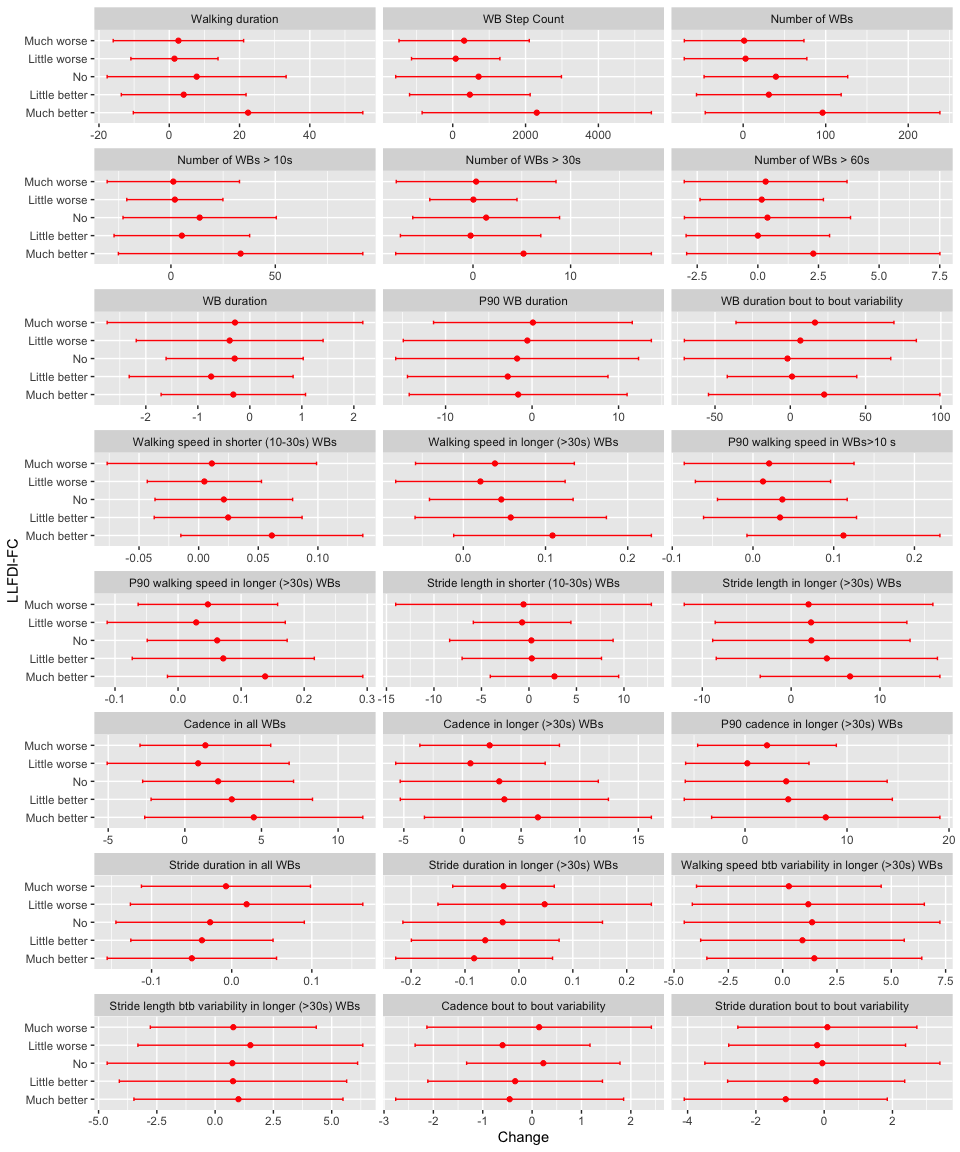

**Figure S2.** Plot of mean changes and standard deviation from first visit to 6-months follow-up, stratified by LLFDI function component categories (acute subgroup n=67 excluded). Btb=Bout to bout

**Table S5**. Change in DMOs from first visit to 6-month follow-up, stratified by SBBP total categories

|  | **SPPB** | **Mean change** | **SD change** | **Effect size** | **n** |
| --- | --- | --- | --- | --- | --- |
| **Walking activity - Volume** | | | | | |
| **Walking duration (min/day)** | | | | | |
|  | Much worse | 1 | 19 | 0.05 | 65 |
|  | No | 9 | 29 | 0.29 | 67 |
|  | Much better | 23 | 29 | 0.80 | 240 |
| **WB Step Count (steps/day)** | | | | | |
|  | Much worse | 142 | 1806 | 0.08 | 65 |
|  | No | 905 | 2736 | 0.33 | 67 |
|  | Much better | 2299 | 2803 | 0.82 | 240 |
| **Walking activity - Pattern** | | | | | |
| **Number of WBs (#/day)** | | | | | |
|  | Much worse | 7 | 87 | 0.08 | 65 |
|  | No | 34 | 107 | 0.32 | 67 |
|  | Much better | 108 | 129 | 0.84 | 240 |
| **Number of WBs > 10s (#/day)** | | | | | |
|  | Much worse | 0 | 30 | -0.02 | 65 |
|  | No | 10 | 50 | 0.21 | 67 |
|  | Much better | 36 | 50 | 0.73 | 240 |
| **Number of WBs > 30s (#/day)** | | | | | |
|  | Much worse | -1 | 7 | -0.19 | 65 |
|  | No | 2 | 11 | 0.17 | 67 |
|  | Much better | 6 | 11 | 0.51 | 240 |
| **Number of WBs > 60s (#/day)** | | | | | |
|  | Much worse | 0 | 4 | -0.02 | 65 |
|  | No | 1 | 4 | 0.23 | 67 |
|  | Much better | 2 | 5 | 0.47 | 240 |
| **WB duration (s)** | | | | | |
|  | Much worse | -0.1 | 1.1 | -0.06 | 65 |
|  | No | -0.7 | 1.8 | -0.35 | 67 |
|  | Much better | -0.4 | 1.6 | -0.24 | 240 |
| **P90 WB duration (s)** | | | | | |
|  | Much worse | -1.1 | 14.7 | -0.07 | 65 |
|  | No | -0.8 | 13.3 | -0.06 | 67 |
|  | Much better | -0.8 | 12.0 | -0.06 | 240 |
| **WB duration bout to bout variability (%)** | | | | | |
|  | Much worse | 5 | 57 | 0.08 | 65 |
|  | No | 9 | 69 | 0.13 | 67 |
|  | Much better | 27 | 72 | 0.38 | 240 |
| **Gait – Pace** | | | | | |
| **Walking speed in shorter (10-30s) WBs (m/s)** | | | | | |
|  | Much worse | 0.02 | 0.06 | 0.40 | 64 |
|  | No | 0.02 | 0.05 | 0.35 | 67 |
|  | Much better | 0.07 | 0.09 | 0.76 | 240 |
| **Walking speed in longer (>30s) WBs (m/s)** | | | | | |
|  | Much worse | 0.04 | 0.10 | 0.46 | 61 |
|  | No | 0.04 | 0.10 | 0.40 | 65 |
|  | Much better | 0.12 | 0.12 | 1.01 | 222 |
| **Maximum walking speed in WBs>10 s (m/s)** | | | | | |
|  | Much worse | 0.03 | 0.09 | 0.36 | 64 |
|  | No | 0.04 | 0.08 | 0.47 | 67 |
|  | Much better | 0.12 | 0.13 | 0.91 | 240 |
| **Maximum walking speed in longer (>30s) WBs (m/s)** | | | | | |
|  | Much worse | 0.04 | 0.12 | 0.34 | 61 |
|  | No | 0.06 | 0.13 | 0.41 | 65 |
|  | Much better | 0.16 | 0.16 | 1.03 | 222 |
| **Stride length in shorter (10-30s) WBs (cm)** | | | | | |
|  | Much worse | 1 | 6 | 0.22 | 64 |
|  | No | 0 | 9 | -0.04 | 67 |
|  | Much better | 3 | 9 | 0.39 | 240 |
| **Stride length in longer (>30s) WBs (cm)** | | | | | |
|  | Much worse | 4 | 10 | 0.36 | 61 |
|  | No | 2 | 11 | 0.17 | 65 |
|  | Much better | 9 | 12 | 0.75 | 222 |
| **Gait – Rhythm** | | | | | |
| **Cadence in all WBs (steps/min)** | | | | | |
|  | Much worse | 1.7 | 4.3 | 0.39 | 65 |
|  | No | 2.3 | 4.7 | 0.49 | 67 |
|  | Much better | 4.9 | 7.4 | 0.66 | 240 |
| **Cadence in longer (>30s) WBs (steps/min)** | | | | | |
|  | Much worse | 1.5 | 7.4 | 0.20 | 61 |
|  | No | 2.5 | 6.9 | 0.37 | 65 |
|  | Much better | 6.8 | 9.5 | 0.71 | 222 |
| **P90 cadence in longer (>30s) WBs (steps/min)** | | | | | |
|  | Much worse | 1.2 | 8.2 | 0.14 | 61 |
|  | No | 3.0 | 7.5 | 0.40 | 65 |
|  | Much better | 9.0 | 11.4 | 0.79 | 222 |
| **Stride duration in all WBs (s)** | | | | | |
|  | Much worse | -0.01 | 0.09 | -0.11 | 65 |
|  | No | -0.01 | 0.09 | -0.16 | 67 |
|  | Much better | -0.07 | 0.14 | -0.46 | 240 |
| **Stride duration in longer (>30s) WBs (s)** | | | | | |
|  | Much worse | 0.00 | 0.16 | -0.01 | 61 |
|  | No | -0.01 | 0.12 | -0.12 | 65 |
|  | Much better | -0.10 | 0.18 | -0.54 | 222 |
| **Gait – Variability** | | | | | |
| **Walking speed bout to bout variability in longer (>30s) WBs (%)** | | | | | |
|  | Much worse | 0 | 6 | 0.04 | 59 |
|  | No | 1 | 4 | 0.20 | 63 |
|  | Much better | 2 | 5 | 0.39 | 205 |
| **Stride length bout to bout variability in longer (>30s) WBs (%)** | | | | | |
|  | Much worse | 0 | 5 | 0.04 | 59 |
|  | No | 1 | 5 | 0.21 | 63 |
|  | Much better | 1 | 5 | 0.28 | 205 |
| **Cadence bout to bout variability (%)** | | | | | |
|  | Much worse | 0 | 2 | 0.02 | 65 |
|  | No | 0 | 2 | -0.09 | 67 |
|  | Much better | -1 | 2 | -0.24 | 240 |
| **Stride duration bout to bout variability (%)** | | | | | |
|  | Much worse | 0 | 3 | -0.02 | 65 |
|  | No | 0 | 2 | -0.04 | 67 |
|  | Much better | -1 | 3 | -0.31 | 240 |

Changers are defined as change in SPPB total according to the established MID, where a difference of at least 1 point for a substantial change in either direction. Please note that it was not possible to categorise those with small changes (0.5 points change according to the literature) on the individual level, since SPPB total only sums up in integer numbers individually.

Colour coding: Green shading shows effect sizes higher than |0.2| and detected changes in the expected direction (light green small effects: |≥0.2 to <0.5|, medium green moderate effects: |≥0.5 to <0.8|, dark green large effects: |≥0.8|). Grey shading shows effect sizes higher than |0.2| and detected changes against expectations. Blue shading shows effect sizes higher than |0.2| in the five bout-to-bout variability DMOs for which we did not have an expectation (with one exception all <0.5). No shading (white) means no effect (effect sizes <0.2).

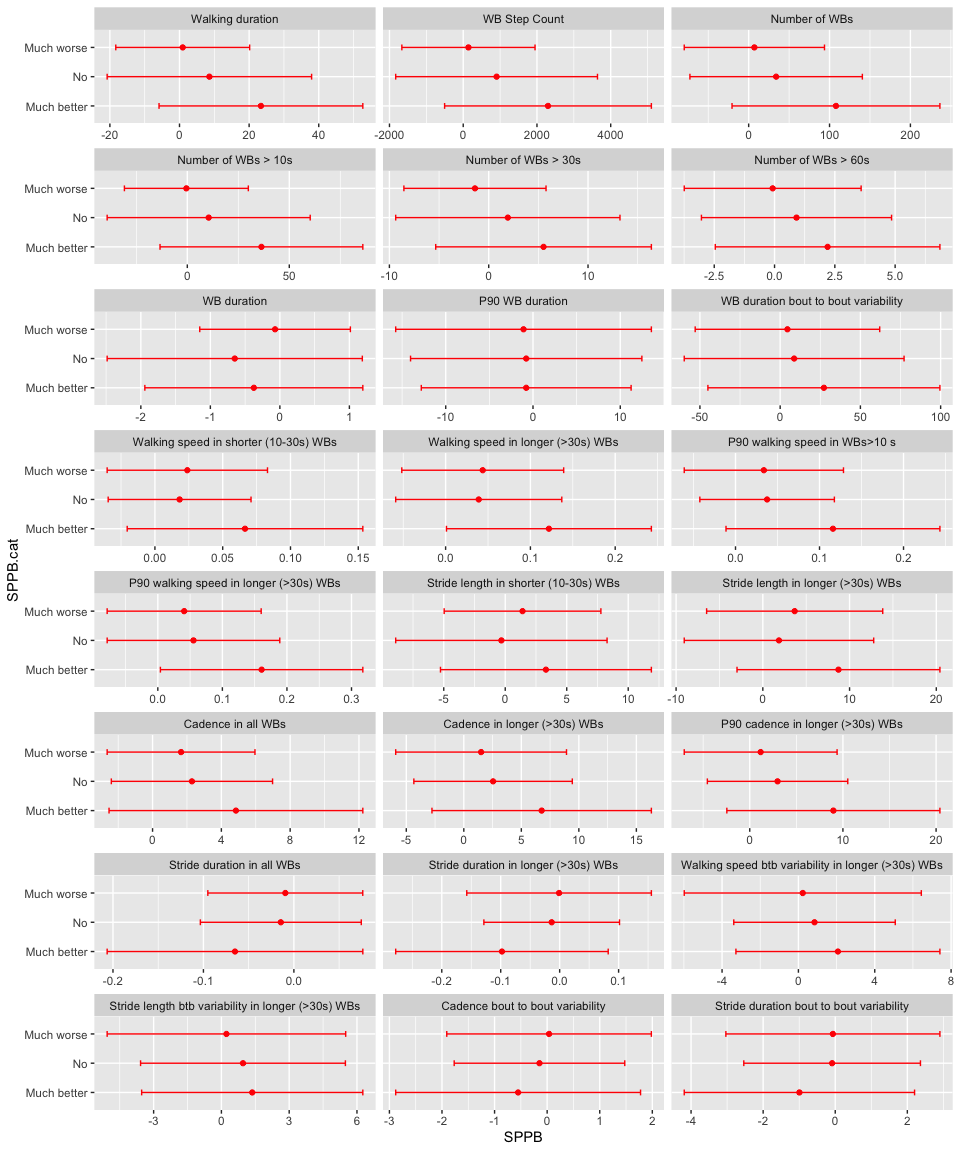

**Figure S3**. Plot of mean changes and standard deviations from first visit to 6-months follow-up, stratified by SPPB total categories. Btb=Bout to bout

**Table S6**. Overview Change in DMOs from first visit to 6-month follow-up, stratified by supervised gait speed (from SPPB) categories

|  | **Gait speed (from SPPB)** | **Mean change** | **SD change** | **Effect size** | **n** |
| --- | --- | --- | --- | --- | --- |
| **Walking activity - Volume** | | | | | |
| **Walking duration (min/day)** | | | | | |
|  | Much worse | 9 | 22 | 0.38 | 44 |
|  | Little worse | 7 | 24 | 0.31 | 26 |
|  | No | 7 | 29 | 0.23 | 62 |
|  | Little better | 13 | 26 | 0.50 | 46 |
|  | Much better | 24 | 30 | 0.80 | 193 |
| **WB Step Count (steps/day)** | | | | | |
|  | Much worse | 8045 | 1938 | 0.42 | 44 |
|  | Little worse | 692 | 2167 | 0.32 | 26 |
|  | No | 646 | 2731 | 0.24 | 62 |
|  | Little better | 1330 | 2455 | 0.54 | 46 |
|  | Much better | 2417 | 2909 | 0.83 | 193 |
| **Walking activity - Pattern** | | | | | |
| **Number of WBs (#/day)** | | | | | |
|  | Much worse | 32 | 84 | 0.38 | 44 |
|  | Little worse | 30 | 124 | 0.25 | 26 |
|  | No | 30 | 109 | 0.28 | 62 |
|  | Little better | 51 | 103 | 0.49 | 46 |
|  | Much better | 115 | 132 | 0.87 | 193 |
| **Number of WBs > 10s (#/day)** | | | | | |
|  | Much worse | 14 | 43 | 0.33 | 44 |
|  | Little worse | 13 | 41 | 0.33 | 26 |
|  | No | 11 | 52 | 0.22 | 62 |
|  | Little better | 14 | 42 | 0.34 | 46 |
|  | Much better | 37 | 50 | 0.73 | 193 |
| **Number of WBs > 30s (#/day)** | | | | | |
|  | Much worse | 3 | 10 | 0.26 | 44 |
|  | Little worse | 1 | 7 | 0.12 | 26 |
|  | No | 1 | 12 | 0.09 | 62 |
|  | Little better | 3 | 10 | 0.33 | 46 |
|  | Much better | 5 | 11 | 0.48 | 193 |
| **Number of WBs > 60s (#/day)** | | | | | |
|  | Much worse | 1 | 4 | 0.33 | 44 |
|  | Little worse | 0 | 3 | 0.07 | 26 |
|  | No | 0 | 4 | 0.06 | 62 |
|  | Little better | 2 | 4 | 0.44 | 46 |
|  | Much better | 2 | 5 | 0.46 | 193 |
| **WB duration (s)** | | | | | |
|  | Much worse | -0.2 | 1.6 | -0.09 | 44 |
|  | Little worse | 0.0 | 1.2 | 0.02 | 26 |
|  | No | -0.2 | 1.3 | -0.17 | 62 |
|  | Little better | -0.6 | 1.5 | -0.43 | 46 |
|  | Much better | -0.5 | 1.7 | -0.27 | 193 |
| **P90 WB duration (s)** | | | | | |
|  | Much worse | -2.1 | 10.6 | -0.19 | 44 |
|  | Little worse | 0.0 | 19.0 | 0.00 | 26 |
|  | No | -1.3 | 14.5 | -0.09 | 62 |
|  | Little better | -0.8 | 10.2 | -0.08 | 46 |
|  | Much better | -0.5 | 12.1 | -0.04 | 193 |
| **WB duration bout to bout variability (%)** | | | | | |
|  | Much worse | 4 | 53 | 0.08 | 44 |
|  | Little worse | 7 | 61 | 0.11 | 26 |
|  | No | 7 | 60 | 0.11 | 62 |
|  | Little better | 9 | 54 | 0.17 | 46 |
|  | Much better | 32 | 79 | 0.41 | 193 |
| **Gait – Pace** | | | | | |
| **Walking speed in shorter (10-30s) WBs (m/s)** | | | | | |
|  | Much worse | 0.01 | 0.05 | 0.21 | 44 |
|  | Little worse | 0.02 | 0.06 | 0.41 | 25 |
|  | No | 0.02 | 0.06 | 0.28 | 62 |
|  | Little better | 0.04 | 0.06 | 0.60 | 46 |
|  | Much better | 0.08 | 0.09 | 0.91 | 193 |
| **Walking speed in longer (>30s) WBs (m/s)** | | | | | |
|  | Much worse | 0.02 | 0.07 | 0.22 | 42 |
|  | Little worse | 0.03 | 0.09 | 0.35 | 25 |
|  | No | 0.05 | 0.10 | 0.44 | 59 |
|  | Little better | 0.09 | 0.10 | 0.87 | 43 |
|  | Much better | 0.14 | 0.12 | 1.15 | 178 |
| **Maximum walking speed in WBs>10 s (m/s)** | | | | | |
|  | Much worse | 0.01 | 0.07 | 0.17 | 44 |
|  | Little worse | 0.03 | 0.09 | 0.38 | 25 |
|  | No | 0.03 | 0.09 | 0.35 | 62 |
|  | Little better | 0.07 | 0.12 | 0.60 | 46 |
|  | Much better | 0.14 | 0.12 | 1.12 | 193 |
| **Maximum walking speed in longer (>30s) WBs (m/s)** | | | | | |
|  | Much worse | 0.02 | 0.09 | 0.21 | 42 |
|  | Little worse | 0.04 | 0.12 | 0.31 | 25 |
|  | No | 0.07 | 0.13 | 0.50 | 59 |
|  | Little better | 0.10 | 0.14 | 0.73 | 43 |
|  | Much better | 0.18 | 0.16 | 1.13 | 178 |
| **Stride length in shorter (10-30s) WBs (cm)** | | | | | |
|  | Much worse | 0 | 5 | 0.04 | 44 |
|  | Little worse | 2 | 4 | 0.40 | 25 |
|  | No | 0 | 8 | -0.02 | 62 |
|  | Little better | 1 | 7 | 0.10 | 46 |
|  | Much better | 4 | 9 | 0.49 | 193 |
| **Stride length in longer (>30s) WBs (cm)** | | | | | |
|  | Much worse | 1 | 8 | 0.17 | 42 |
|  | Little worse | 3 | 7 | 0.40 | 25 |
|  | No | 3 | 10 | 0.26 | 59 |
|  | Little better | 6 | 11 | 0.61 | 43 |
|  | Much better | 10 | 12 | 0.81 | 178 |
| **Gait – Rhythm** | | | | | |
| **Cadence in all WBs (steps/min)** | | | | | |
|  | Much worse | 1.0 | 3.3 | 0.30 | 44 |
|  | Little worse | 1.7 | 5.6 | 0.30 | 26 |
|  | No | 1.9 | 5.0 | 0.39 | 62 |
|  | Little better | 3.5 | 5.2 | 0.68 | 46 |
|  | Much better | 5.4 | 7.6 | 0.71 | 193 |
| **Cadence in longer (>30s) WBs (steps/min)** | | | | | |
|  | Much worse | 0.8 | 5.8 | 0.13 | 42 |
|  | Little worse | 1.2 | 8.3 | 0.14 | 25 |
|  | No | 2.9 | 7.4 | 0.40 | 59 |
|  | Little better | 4.4 | 9.3 | 0.47 | 43 |
|  | Much better | 7.5 | 9.5 | 0.79 | 178 |
| **P90 cadence in longer (>30s) WBs (steps/min)** | | | | | |
|  | Much worse | 0.6 | 6.2 | 0.09 | 42 |
|  | Little worse | 1.3 | 8.8 | 0.15 | 25 |
|  | No | 3.6 | 9.0 | 0.40 | 59 |
|  | Little better | 5.0 | 10.4 | 0.48 | 43 |
|  | Much better | 9.9 | 11.4 | 0.87 | 178 |
| **Stride duration in all WBs (s)** | | | | | |
|  | Much worse | 0.00 | 0.07 | -0.04 | 44 |
|  | Little worse | -0.02 | 0.10 | -0.18 | 26 |
|  | No | -0.01 | 0.10 | -0.13 | 62 |
|  | Little better | -0.04 | 0.08 | -0.55 | 46 |
|  | Much better | -0.07 | 0.15 | -0.48 | 193 |
| **Stride duration in longer (>30s) WBs (s)** | | | | | |
|  | Much worse | 0.01 | 0.11 | 0.08 | 42 |
|  | Little worse | -0.01 | 0.13 | -0.04 | 25 |
|  | No | -0.01 | 0.12 | -0.05 | 59 |
|  | Little better | -0.05 | 0.19 | -0.28 | 43 |
|  | Much better | -0.11 | 0.18 | -0.62 | 178 |
| **Gait – Variability** | | | | | |
| **Walking speed bout to bout variability in longer (>30s) WBs (%)** | | | | | |
|  | Much worse | 0 | 5 | -0.01 | 41 |
|  | Little worse | 1 | 5 | 0.18 | 25 |
|  | No | 2 | 6 | 0.35 | 54 |
|  | Little better | 0 | 4 | -0.05 | 43 |
|  | Much better | 2 | 5 | 0.42 | 163 |
| **Stride length bout to bout variability in longer (>30s) WBs (%)** | | | | | |
|  | Much worse | 0 | 4 | 0.09 | 41 |
|  | Little worse | 1 | 5 | 0.22 | 25 |
|  | No | 1 | 5 | 0.17 | 54 |
|  | Little better | 1 | 4 | 0.12 | 43 |
|  | Much better | 1 | 5 | 0.29 | 163 |
| **Cadence bout to bout variability (%)** | | | | | |
|  | Much worse | 0 | 2 | 0.06 | 44 |
|  | Little worse | 0 | 1 | -0.05 | 26 |
|  | No | 0 | 1 | -0.09 | 62 |
|  | Little better | -1 | 2 | -0.33 | 46 |
|  | Much better | -1 | 2 | -0.23 | 193 |
| **Stride duration bout to bout variability (%)** | | | | | |
|  | Much worse | 0 | 2 | -0.04 | 44 |
|  | Little worse | 0 | 3 | -0.11 | 26 |
|  | No | -1 | 3 | -0.20 | 62 |
|  | Little better | 0 | 3 | -0.02 | 46 |
|  | Much better | -1 | 3 | -0.31 | 193 |

Changers are defined as change in supervised gait speed according to the established MID, where the MID of 0.05 m/s denoted a small change (little worse, little better) and 0.1 m/s substantial change (much worse, much better) in either direction.

Colour coding: Green shading shows effect sizes higher than |0.2| and detected changes in the expected direction (light green small effects: |≥0.2 to <0.5|, medium green moderate effects: |≥0.5 to <0.8|, dark green large effects: |≥0.8|). Grey shading shows effect sizes higher than |0.2| and detected changes against expectations. Blue shading shows effect sizes higher than |0.2| in the five bout-to-bout variability DMOs for which we did not have an expectation (with one exception all <0.5). No shading (white) means no effect (effect sizes <0.2).

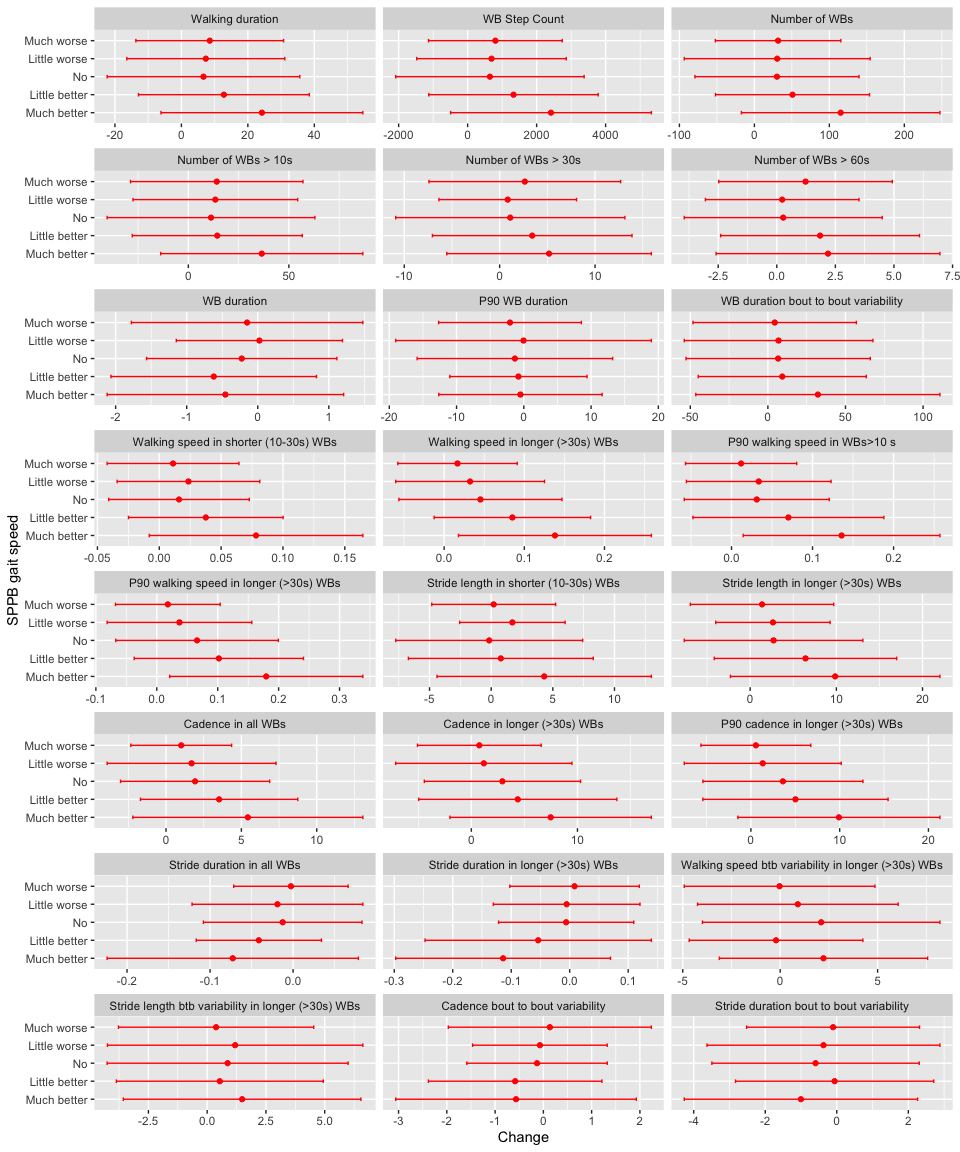

**Figure S4**. Plot of mean changes and standard deviations from first visit to 6-months follow-up, stratified by supervised gait speed (from SPPB) categories. Btb=Bout to bout

Table S7 summarizes all colour coded standardized mean changes of the DMOs from the Tables S3-S6 and provides an overview of all results per DMO.

**Table S7**. Summary of all standardized mean changes (effect sizes) per DMO, as reported in Tables S3-S6

| **DMOs** | **GIC** | **LLFDI-FC^1)^** | **SPPB total** | **Gait speed** |
| --- | --- | --- | --- | --- |
| **n** | 11: much worse  26: little worse  49: no change  105: little better  126: much better | 28: Much worse  24: little worse  62: no change  56: little better  144: much better | 65: much worse  67: no change  240: much better | 44: much worse  26: little worse  62: no change  46: little better  193: much better |
| **Walking activity - Volume** | |  |  |  |
| Walking duration (min/day) | -0.17; 0.77; 0.38; 0.47; 0.90 | 0.14; 0.10; 0.29; 0.19; 0.69 | 0.05; 0.29; 0.80 | 0.38; 0.31; 0.23; 0.50; 0.80 |
| WB Step Count (#/day) | -0.27; 0.78; 0.40; 0.50; 0.91 | 0.18; 0.05; 0.29; 0.25; 0.74 | 0.08; 0.33; 0.82 | 0.42; 0.32; 0.24; 0.54; 0.83 |
| **Walking activity – Pattern** | |  |  |  |
| Number of WBs (#/day) | -0.34; 0.82; 0.41; 0.50; 0.90 | 0.02; 0.02; 0.51; 0.33; 0.69 | 0.08; 0.32; 0.84 | 0.38; 0.25; 0.28; 0.49; 0.87 |
| Number of WBs > 10s (#/day) | -0.23; 0.69; 0.34; 0.34; 0.86 | 0.04; 0.06; 0.39; 0.14; 0.57 | -0.02; 0.21; 0.73 | 0.33; 0.33; 0.22; 0.34; 0.73 |
| Number of WBs > 30s (#/day) | 0.16; 0.19; 0.20; 0.22; 0.64 | 0.04; -0.02; 0.14; -0.07; 0.41 | -0.19; 0.17; 0.51 | 0.26; 0.12; 0.09; 0.33; 0.48 |
| Number of WBs > 60s (#/day) | 0.65; 0.27; 0.10; 0.26; 0.60 | 0.10; 0.06; 0.08; -0.09; 0.45 | -0.02; 0.23; 0.47 | 0.33; 0.07; 0.06; 0.44; 0.46 |
| WB duration (s) | -0.13; -0.32; -0.25; -0.23; -0.18 | -0.12; -0.25; -0.24; -0.45; -0.20 | -0.06; -0.35; -0.24 | -0.09; 0.02; -0.17; -0.43; -0.27 |
| P90 WB duration (s) | -0.08; -0.27; -0.08; -0.09; 0.09 | 0.01; -0.04; -0.13; -0.23; -0.10 | -0.07; -0.06; -0.06 | -0.19; 0.00; -0.09; -0.08; -0.04 |
| WB duration bout to bout variability (%) | 0.18; 0.35; 0.07; 0.29; 0.36 | 0.31; 0.08; -0.06; -0.05; 0.29 | 0.08; 0.13; 0.38 | 0.08; 0.11; 0.11; 0.17; 0.41 |
| **Gait – Pace** | |  |  |  |
| Walking speed in shorter (10-30s) WBs (m/s) | -0.36; 0.69; 0.21; 0.56; 1.01 | 0.13; 0.09; 0.34; 0.40; 0.79 | 0.40; 0.35; 0.76 | 0.21; 0.41; 0.28; 0.60; 0.91 |
| Walking speed in longer (>30s) WBs (m/s) | -0.45; 0.51; 0.47; 0.76; 1.05 | 0.40; 0.21; 0.52; 0.47; 0.90 | 0.46; 0.40; 1.01 | 0.22; 0.35; 0.44; 0.87; 1.15 |
| P90 walking speed in WBs>10 s (m/s) | -0.27; 0.63; 0.36; 0.68; 1.04 | 0.19; 0.13; 0.43; 0.33; 0.94 | 0.36; 0.47; 0.91 | 0.17; 0.38; 0.35; 0.60; 1.12 |
| P90 walking speed in longer (>30s) WBs (m/s) | -0.38; 0.51; 0.53; 0.75; 1.05 | 0.43; 0.21; 0.56; 0.47; 0.89 | 0.34; 0.41; 1.03 | 0.21; 0.31; 0.50; 0.73; 1.13 |
| Stride length in shorter (10-30s) WBs (cm) | -0.26; 0.50; -0.16; 0.25; 0.66 | -0.04; -0.18; -0.01; 0.09; 0.39 | 0.22; -0.04; 0.39 | 0.04; 0.40; -0.02; 0.10; 0.49 |
| Stride length in longer (>30s) WBs (cm) | -0.32; 0.26; 0.21; 0.49; 0.85 | 0.14; 0.20; 0.18; 0.32; 0.64 | 0.36; 0.17; 0.75 | 0.17; 0.40; 0.26; 0.61; 0.81 |
| **Gait – Rhythm** | |  |  |  |
| Cadence in all WBs (steps/min) | -0.27; 0.51; 0.51; 0.55; 0.67 | 0.31; 0.16; 0.43; 0.59; 0.61 | 0.39; 0.49; 0.66 | 0.30; 0.30; 0.39; 0.68; 0.71 |
| Cadence in longer (>30s) WBs (steps/min) | -0.27; 0.53; 0.52; 0.60; 0.73 | 0.39; 0.14; 0.35; 0.39; 0.65 | 0.20; 0.37; 0.71 | 0.13; 0.14; 0.40; 0.47; 0.79 |
| P90 cadence in longer (>30s) WBs (steps/min) | -0.32; 0.51; 0.54; 0.65; 0.80 | 0.32; 0.07; 0.38; 0.41; 0.69 | 0.14; 0.40; 0.79 | 0.09; 0.15; 0.41; 0.48; 0.87 |
| Stride duration in all WBs (s) | 0.26; -0.38; -0.45; -0.34; -0.38 | -0.07; 0.14; -0.23; -0.39; -0.45 | -0.11; -0.16; -0.46 | -0.04; -0.18; -0.13; -0.55; -0.48 |
| Stride duration in longer (>30s) WBs (s) | 0.12; -0.54; -0.43; -0.34; -0.49 | -0.30; 0.22; -0.15; -0.43; -0.55 | -0.01; -0.12; -0.54 | 0.08; -0.04; -0.05; -0.28; -0.62 |
| **Gait – Variability** | |  |  |  |
| Walking speed bout to bout variability in longer (>30s) WBs (%) | 0.07; 0.13; 0.33; 0.26; 0.37 | 0.07; 0.21; 0.21; 0.22; 0.28 | 0.04; 0.20; 0.39 | -0.01; 0.18; 0.35; -0.05; 0.42 |
| Stride length bout to bout variability in longer (>30s) WBs (%) | 0.54; 0.03; 0.01; 0.28; 0.19 | 0.22; 0.30; 0.12; 0.15; 0.20 | 0.04; 0.21; 0.28 | 0.09; 0.22; 0.17; 0.12; 0.29 |
| Cadence bout to bout variability (%) | 0.13; -0.33; 0.06; -0.26; -0.20 | 0.06; -0.41; 0.21; -0.20; -0.21 | 0.02; -0.09; -0.24 | 0.06; -0.05; -0.09; -0.33; -0.23 |
| Stride duration bout to bout variability (%) | 0.04; -0.29; 0.20; -0.17; -0.41 | 0.04; -0.11; 0.02; -0.08; -0.38 | -0.02; -0.04; -0.31 | -0.04; -0.11; -0.20; -0.02; -0.31 |

^1)^Acute patients excluded (n=67)

Changers are defined as exceeding established MIDs: *LLFDI function component*: |≥2| = small change (little worse, little better), |≥5| = substantial change (much worse, much better); *SPPB total*: |≥2| = substantial change; *supervised gait speed*: |≥0.05| = small change (little worse, little better), |≥0.1| = substantial change (much worse, much better).

Colour coding: Green shading shows effect sizes larger than |0.2| and detected changes in the expected direction (light green small effects: |≥0.2 to <0.5|, medium green moderate effects: |≥0.5 to <0.8|, dark green large effects: |≥0.8|). Grey shading shows effect sizes higher than |0.2| and detected changes against expectations. Blue shading shows effect sizes higher than |0.2| in the five bout-to-bout variability DMOs for which we did not have an expectation (with one exception all <0.5). No shading (white) means no effect (effect sizes <0.2).

Seventeen out of 24 available DMOs showed at least moderate positive effects in the expected direction of the respective anchors:

***Moderate effects (0.5-0.8, n=7)***: Number of WBs > 30s, Number of WBs > 60s, Stride length in shorter (10-30s) WBs, Cadence in all WB, Cadence in longer (>30s) WBs, Stride duration in all WBs, Stride duration in longer (>30s) WBs

***Large effects (>0.8, n=10)****:* Walking duration, WB Step Count, Number of WBs, Number of WBs > 10s, Walking speed in shorter (10-30s) WBs, Walking speed in longer (>30s) WBs, P90 walking speed in WB > 10 s, P90 walking speed in longer (>30s) WB, Stride length in longer (>30s) WBs, P90 cadence in longer (>30s) WB

***No or only small effects (0-0.5, n=7)***: WB duration, P90 WB duration, WB duration bout to bout variability, Walking speed bout to bout variability in longer (>30s) WBs, Stride length bout to bout variability in longer (>30s) WBs (one exception >0.5), Cadence bout to bout variability, Stride duration bout to bout variability

**Minimal important difference**

Table S8 shows the correlation between the change scores of the clinical outcomes and the change scores of the DMOs. Correlations large enough (>0.3) to be used for the anchor-based method are shaded in green.

**Table S8**. Spearman’s correlations between change in anchors and change in DMOs

| **DMOs** | **LLFDI-FC**^1)^ | **SPPB** | **Supervised gait speed** |
| --- | --- | --- | --- |
| **Walking activity - Volume** |  |  |  |
| Walking duration (min/day) | 0.33 | 0.47 | 0.39 |
| WB Step Count (steps/day) | 0.35 | 0.48 | 0.41 |
| **Walking activity – Pattern** |  |  |  |
| Number of WBs (#/day) | 0.34 | 0.52 | 0.44 |
| Number of WBs > 10s (#/day) | 0.30 | 0.46 | 0.38 |
| Number of WBs > 30s (#/day) | 0.22 | 0.36 | 0.25 |
| Number of WBs > 60s (#/day) | 0.20 | 0.31 | 0.23 |
| WB duration (s) | 0.03 | -0.06 | -0.06 |
| P90 WB duration (s) | 0.04 | 0.07 | 0.04 |
| WB duration bout to bout variability (%) | 0.15 | 0.28 | 0.27 |
| **Gait - Pace** |  |  |  |
| Walking speed in shorter (10-30s) WBs (m/s) | 0.33 | 0.37 | 0.44 |
| Walking speed in longer (>30s) WBs (m/s) | 0.32 | 0.43 | 0.51 |
| Maximum walking speed in WBs>10 s (m/s) | 0.40 | 0.45 | 0.52 |
| Maximum walking speed in longer (>30s) WBs (m/s) | 0.31 | 0.47 | 0.52 |
| Stride length in shorter (10-30s) WBs (cm) | 0.18 | 0.21 | 0.26 |
| Stride length in longer (>30s) WBs (cm) | 0.20 | 0.36 | 0.37 |
| **Gait - Rhythm** |  |  |  |
| Cadence in all WBs (steps/min) | 0.25 | 0.30 | 0.33 |
| Cadence in longer (>30s) WBs (steps/min) | 0.27 | 0.33 | 0.40 |
| Maximum cadence in longer (>30s) WBs (steps/min) | 0.28 | 0.39 | 0.45 |
| Stride duration in all WBs (s) | -0.17 | -0.31 | -0.30 |
| Stride duration in longer (>30s) WBs (s) | -0.20 | -0.31 | -0.41 |
| **Gait - Variability** |  |  |  |
| Walking speed bout to bout variability in longer (>30s) WBs (%) | 0.11 | 0.20 | 0.16 |
| Stride length bout to bout variability in longer (>30s) WBs | 0.05 | 0.20 | 0.13 |
| Cadence bout to bout variability (%) | -0.06 | -0.12 | -0.13 |
| Stride duration bout to bout variability (%) | -0.16 | -0.14 | -0.16 |

1) Subgroup without acute patients (n=67 excluded).

Table S9 summarizes the AUC of the change scores of each DMO-anchor pair, shaded in green if AUC was large enough (≥0.7)

**Table S9**. Area under the curve values for binary anchors and change in DMOs

| **DMOs** | **GIC overall** | **LLFDI-FC**^1)^ | **SPPB** | **Supervised gait speed** |
| --- | --- | --- | --- | --- |
| **Walking activity - Volume** |  |  |  |  |
| Walking duration (min/day) | 0.62 | 0.58 | 0.70 | 0.68 |
| WB Step Count (steps/day) | 0.63 | 0.61 | 0.71 | 0.69 |
| **Walking activity – Pattern** |  |  |  |  |
| Number of WBs (#/day) | 0.62 | 0.59 | 0.72 | 0.71 |
| Number of WBs > 10s (#/day) | 0.61 | 0.56 | 0.71 | 0.68 |
| Number of WBs > 30s (#/day) | 0.60 | 0.56 | 0.67 | 0.61 |
| Number of WBs > 60s (#/day) | 0.59 | 0.57 | 0.63 | 0.59 |
| WB duration (s) | 0.51 | 0.50 | 0.51 | 0.47 |
| P90 WB duration (s) | 0.56 | 0.55 | 0.56 | 0.49 |
| WB duration bout to bout variability (%) | 0.58 | 0.55 | 0.61 | 0.61 |
| **Gait - Pace** |  |  |  |  |
| Walking speed in shorter (10-30s) WBs (m/s) | 0.68 | 0.63 | 0.66 | 0.72 |
| Walking speed in longer (>30s) WBs (m/s) | 0.66 | 0.64 | 0.69 | 0.73 |
| Maximum walking speed in WBs>10 s (m/s) | 0.69 | 0.65 | 0.69 | 0.76 |
| Maximum walking speed in longer (>30s) WBs (m/s) | 0.66 | 0.64 | 0.71 | 0.73 |
| Stride length in shorter (10-30s) WBs (cm) | 0.61 | 0.54 | 0.58 | 0.64 |
| Stride length in longer (>30s) WBs (cm) | 0.64 | 0.58 | 0.65 | 0.66 |
| **Gait - Rhythm** |  |  |  |  |
| Cadence in all WBs (steps/min) | 0.59 | 0.60 | 0.62 | 0.65 |
| Cadence in longer (>30s) WBs (steps/min) | 0.61 | 0.63 | 0.66 | 0.67 |
| P90 cadence in longer (>30s) WBs (steps/min) | 0.62 | 0.64 | 0.68 | 0.69 |
| Stride duration in all WBs (s) | 0.54 | 0.46 | 0.64 | 0.64 |
| Stride duration in longer (>30s) WBs (s) | 0.55 | 0.56 | 0.66 | 0.68 |
| **Gait - Variability** |  |  |  |  |
| Walking speed bout to bout variability in longer (>30s) WBs (%) | 0.54 | 0.55 | 0.59 | 0.59 |
| Stride length bout to bout variability in longer (>30s) WBs (%) | 0.56 | 0.54 | 0.58 | 0.58 |
| Cadence bout to bout variability (%) | 0.55 | 0.51 | 0.56 | 0.53 |
| Stride duration bout to bout variability (%) | 0.58 | 0.57 | 0.58 | 0.57 |

1) Subgroup without acute patients (n=67 excluded).
